# The Natural History of Untreated Pulmonary Tuberculosis in Adults: A Systematic Review and Meta-Analysis

**DOI:** 10.1101/2022.08.30.22279374

**Authors:** Bianca Sossen, Alexandra Richards, Torben Heinsohn, Beatrice Frascella, Federica Balzarini, Aurea Oradini-Alacreu, Anna Odone, Ewelina Rogozińska, Brit Häcker, Frank Cobelens, Katharina Kranzer, Rein MGJ Houben, Hanif Esmail

## Abstract

**BACKGROUND:** Key stages in TB disease can be delineated by radiology, microbiology and symptoms, but transition between relevant stages remains unclear. We sought to quantify progression and regression across the spectrum of TB disease by systematically reviewing studies of individuals with untreated TB undergoing follow up.

**METHODS:** We searched PubMED, EMBASE and Web of Science until December 31^st^ 1960, the Index Medicus between 1895 and 1945, and extensive investigator collections without date restriction - in English and German. Eligible studies were observational cohorts and clinical trials, presenting adults/adolescents with TB or recent TB exposure, undergoing follow-up for at least 12 months without therapeutic intervention. Two authors independently reviewed titles/abstracts and full texts for inclusion. Quality was assessed with a modified Newcastle-Ottawa Score, excluding highly biased studies. Summary estimates were extracted to align with TB disease transitions in a conceptual model, and we used meta-analysis of proportions with random-effects to synthesise the extracted data. This study is registered with PROSPERO (CRD42019152585).

**FINDINGS:** 10477 titles were screened and 1648 full texts reviewed. 223 met inclusion criteria. 109 were excluded for high risk of bias and 90 did not have extractable data. 24 studies (34 cohorts) were included. Progression from microbiologically negative to positive disease in those with radiographic TB evidence occurred at an annualized rate of 9.71% (95% CI:6.17-13.34) with “active” TB imaging, and 1.06% (95% CI:0.31-1.82) with “inactive” TB imaging. Reversion from microbiologically-positive to -undetectable in prospective cohorts occurred at an annualized rate of 12.40% (95% CI: 6.81-17.99). Studies reported symptoms poorly not allowing for direct estimation of transitions for subclinical (asymptomatic, culture positive) disease.

**INTERPRETATION:** We present the risk of progression in those with radiographic evidence of disease and the rate of self-cure for microbiologically positive disease to inform global disease burden estimates, clinical guidelines and policy decisions.

## INTRODUCTION

Despite a clinical awareness of tuberculosis (TB) for centuries, its natural history is incompletely understood. We have oscillated between characterizing TB with binary states of latent infection and active disease, to a condition existing on a dynamic continuum(1–4). In the early 20th century, TB control relied on early identification of those with evidence of disease, particularly through chest X-ray (CXR) screening. Researchers were able to highlight the heterogeneity and dynamics of disease evolution between individuals, through longitudinal assessment(5–8). With the discovery of effective treatment in the mid-20^th^ century and driven by the need for scalable, programmatic treatment algorithms, a binary description of disease states reflecting two extremes (‘latent infection’ and ‘active disease’) became established(9). Although this provided a useful paradigm, the more nuanced understanding of disease natural history was arguably forgotten.

An accurate understanding of the kinetics of TB natural history is now increasingly critical at both population and individual level, with implications for disease management, population-level prevention and control, and disease burden estimations. Treatment of patients that fall between active and latent TB - for instance having abnormalities suggestive of active disease on X-ray but microbiologically negative - is not adequately covered by management algorithms, but progress could be driven by adequate understanding of the risk of disease progression. A better understanding of this natural history is also a key priority for vaccine development(10). In addition, estimates of TB burden currently rely strongly on assumptions around the progression, regression and mortality from untreated TB, of which only mortality estimates are informed by systematic review of available literature(11–13). Furthermore, estimation methods do not cater to different stages of TB which are detected in disease prevalence surveys, including individuals who have culture positive disease but a negative symptom screen (referred to as subclinical), or those with TB suggestive X-rays(14). Given the implications for health care seeking and potential for interrupting or preventing transmission, a better understanding of this natural history is key to inform TB burden estimation and policies for care and prevention.

Within the disease continuum, key stages in the evolution of pulmonary TB can be marked by diagnostic tests that have been available for over a century, to allow for categorization within a widely accepted conceptual framework (Figure 1)(1,2). The emergence of disease pathology is first visible by typical radiographic features. Microbiological detection in sputum signals presence of bacilli (and potential infectiousness), and the reporting of symptoms marks the development of active, clinical disease. Transitions across all of these stages can only be fully studied in the absence of treatment and hence can no longer be ethically investigated. We conducted a systematic review focusing on articles from the pre-chemotherapy era to determine which of the transitions could be adequately described by existing literature, with the aim of providing parameters for the rate of progression and regression of disease across the spectrum.

**Figure 1:**
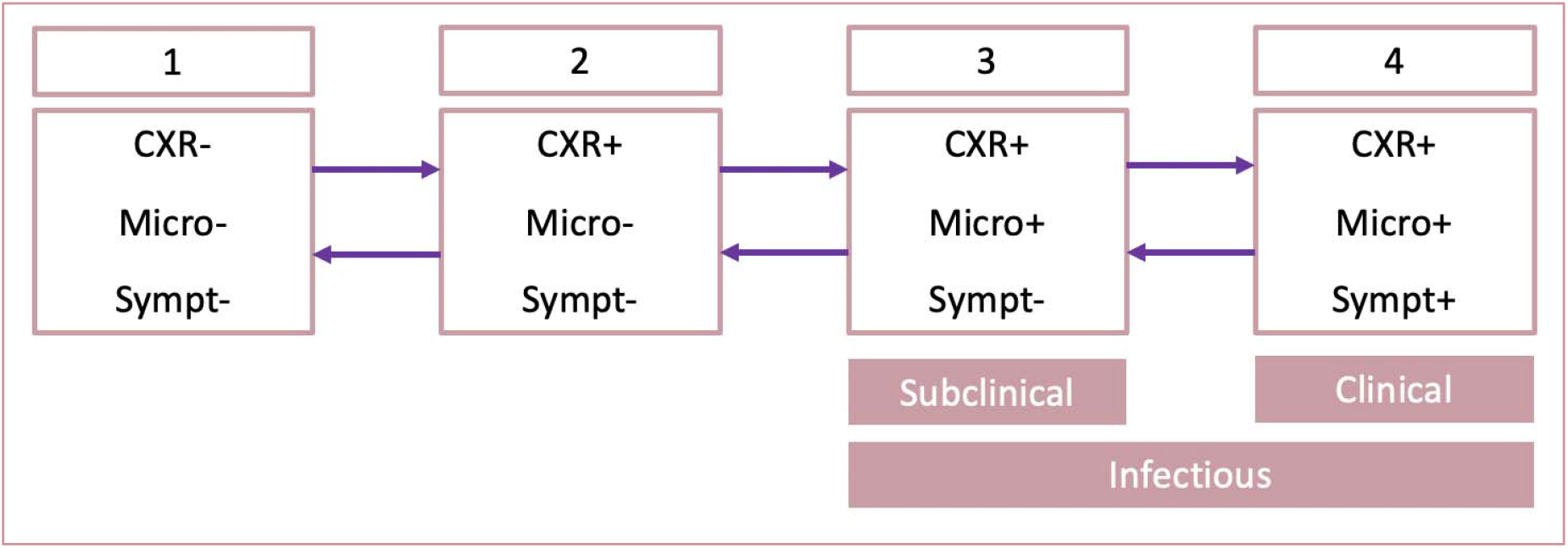
Conceptual framework of transitions occurring in the natural history of tuberculosis. The design of this conceptual framework is based on the available literature regarding the natural history of TB, where a subclinical group is included(1,15,16). The figure demonstrates that individuals would undulate between states of having (1) normal chest x-ray, negative microbiology and being asymptomatic, to (2) chest x-ray abnormalities, but still having negative microbiology and being asymptomatic, to (3) chest x-ray abnormalities with positive microbiology but being asymptomatic, to (4) chest x-ray abnormalities with positive microbiology and being symptomatic. We recognize individuals do not always fall into these groupings while transitioning along the spectrum of disease, for example an individual may present with an abnormal chest X-ray and symptoms that may represent TB but have negative microbiology. We have made allowances to capture all combinations of CXR, microbiology and symptoms status within the review.

CXR=Chest X-ray; Micro=Microbiology; Sympt=Symptoms

#### Panel: Research in context

##### Evidence before this study

Certain aspects of the natural history of TB in the absence of treatment have been previously estimated through systematic review of the literature. Several authors have assessed the progression of latent infection (defined by TST or Interferon-gamma releasing assay (IGRA)) to active disease (usually defined by bacteriological positivity). However, these studies took a binary approach to categorizing TB, not recognizing the spectrum of disease states. Tiemersma *et al* took a systematic approach to determine the mortality of untreated TB disease distinguishing the outcome of ‘open’ and ‘closed’ disease, interpreted as smear positive and smear negative disease respectively. However, they did not focus on the dynamics of progression and regression of early stages of disease. Marais *et al* have investigated the natural history of TB in childhood, focusing on age specific rates and nature of disease progression following primary infection in children. Key gaps in our understanding of disease natural history therefore remain to be systematically assessed. Most notably, in individuals with changes consistent with early TB on CXR (as found on national prevalence surveys), the risk of progression to bacteriologically positive disease has not been sufficiently summarized, nor do we know the progression and regression from bacteriologically positive, symptom screen negative disease, or adequately understand regression (or ‘self-cure) from classic ‘active disease’, i.e. bacteriologically positive, symptom screen positive.

##### Added value of this study

This systematic review has used the enormous body of historical literature to better capture progression and regression across the spectrum of TB disease stages: defined by chest imaging, sputum microbiology and symptom status. We identified 34 cohorts with a combined sample of 139,212 participants that contributed to our analysis. We show progression to bacteriologically positive disease of 10%/year from individuals with CXRs suggestive of active TB, and quantify reversion (self-cure) from microbiologically-positive disease, but could not directly quantify progression into and regression from subclinical disease

##### Implications of all the available evidence

With the currently available and newly generated evidence, we have a better understanding of the kinetics of TB natural history. High progression from CXR positive disease calls for a reconsideration of treatment guidelines. In addition, the comprehensive data collected could enable modelling to inform kinetics around subclinical disease as well. Finally, this improved understanding will allow for efforts to be refined when attempting to study and improve diagnostic and prognostic biomarkers, and likely allow for greater benefit to be derived from targeted TB prevention and treatment efforts.

## METHODS

### Search strategy and selection criteria

This systematic review and meta-analysis was conducted following a protocol registered at PROSPERO (CRD42019152585). The study is reported in accordance with the Preferred Reporting Items for Systematic Reviews and Meta-analyses (PRIMSA) guidelines(17). We searched for articles from the pre-chemotherapy era combining electronic and manual searches. Electronic searches were conducted in Medline (via PubMED), EMBASE and Web of Science from the start of the database (1946, 1947, and 1900 respectively) until 31^st^ December 1960, in two languages with high yield for study designs of interest in this period: English and German. Additionally, we manually searched titles from Index Medicus between 1903 and 1945; volumes from 1895-1902 were not available. The systematic search was restricted to manuscripts published prior to 1960 to include cohorts observed from the pre-chemotherapy era while allowing for a publication delay of earlier cohorts. Furthermore, supplementary searches were conducted in extensive author collections. Further references were snowballed from those articles that met the criteria for data extraction and from key review articles. Personal libraries and snowballed references were searched without date restriction.

Electronic search terms used both modern and historical terminology in English and German (full search strategies in supplementary appendix). All titles were imported into Covidence systematic review software (Veritas Health Innovation, Melbourne, Australia). After de-duplication, titles and abstracts were screened for relevance by two independent reviewers, with a third reviewer resolving conflicts. Full text articles were sought online, within the library stores at the Wellcome and British libraries (English articles) and the library of the German Central Committee against Tuberculosis (DZK) and the German Tuberculosis Archive (DTA) (German articles), and on online archive websites (e.g HathiTrust.org and archive.org). If manuscripts could not be found through any of these sources, they were not included. At full-text stage, two independent reviewers reviewed eligibility. Articles were included if they presented a longitudinal cohort of at least 25 adolescents (≥10 years) and/or adults followed up (radiologically, microbiologically and clinically) for at least 12 months from the point of either (1) positive TST following recent TB exposure, (2) radiographic changes suggestive of TB or (3) positive microbiology for TB (smear microscopy and/or mycobacterial culture). A minimum of 12 months was selected in order to ensure an adequate number of events. Articles were excluded if they made no attempt at microbiological confirmation of disease, presented no new data (i.e. review article), all participants received a therapeutic (medical or surgical) intervention or those who did not receive a therapeutic intervention could not have data extracted separately, or where ≥5% of the cohort were paediatric (<10 years) and these children could not be separated from the adolescent/adult data.

Eligible articles were assessed for risk of bias with an adapted Newcastle-Ottawa Scale (NOS) to a maximum of seven stars (NOS - General Quality Assessment) by two reviewers per language (supplementary table 1) with conflicts resolved by consensus. To pass the quality assessment, studies could only lose two stars in the “Study Selection” and “Outcome” domains of the NOS. The “comparability” domain was not assessed as this systematic review did not use control groups. An additional quality assessment tool was designed to assess the quality of specific diagnostic compartments in study cohorts i.e. radiological, microbiological and symptoms (supplementary table 1). While this Specific Quality Assessment was captured to get a sense of quality of the study designs, it did not inform study eligibility. Those that passed the NOS were extracted in a standardized electronic tool by one reviewer and then datapoints confirmed by a second reviewer with conflicts were resolved by consensus, involving input from additional reviewers if needed.

### Data extraction and analysis

We extracted data corresponding to the proportion of individuals in the cohort transitioning between diagnostic states (figure 1) over a specified period of time. Recognizing that description of symptom status in particular may not always be explicit by current standards this could be recorded as unknown as long as microbiological status was clear. Where authors differentiated abnormal chest imaging that was suggestive of TB versus not suggestive then we only extracted the TB-suggestive group as abnormal. In addition, where authors provided a subgroup of abnormal chest x-rays that were limited to only calcified nodules then we did not deem these to be an abnormal x-ray for the purpose of this review. The clinical classification method of the National Tuberculosis Association Diagnostic Standards and Classification of Tuberculosis; facilitated extraction of the data(18).

Certain studies presented the proportion of individuals who progressed within a window of time rather than a specific time point; in these cases, we have presented datapoints as at the midpoint of the time window provided. All summary estimates are presented with 95% confidence intervals, calculated from the point data provided. To allow for exploration of the data and any heterogeneity, we attempted to collect data on variables of interest, namely: age distribution, sex, frequency of follow up visits, microbiological test used (i.e. culture versus smear), CXR characteristics described by the historical study’s authors, TST data, local disease burden as per today’s WHO classification(19), features of the study design (i.e. passive versus active versus mixed case finding and whether the data was generated from two cross-sectional assessments of participants (“single follow-up”) or through a cumulative count of events over time (“cumulative count”)), the enrollment setting, and symptom status.

To allow comparison of the varying follow-up times, the last data point of each study was annualised and the expected number transitioning in the first year calculated. The variance of the annualised rate was then calculated using the escalc function from the metafor package(20), specifying the raw proportion measure. Meta-analysis was then conducted using the rma function with the study outcome and variance as inputs. By default each study was weighted proportional to the inverse of the variance calculated in the previous step. The forest plots were created using the forest function from the meta package. Confidence interval proportions were limited to between 0 and 1 by the observation limit argument within the forest function. Sub analyses were also conducted using the rma function and added to the forest plot using the addpoly function from metafor. Heterogeneity was assessed with the I^2^ and tau^2^ statistics.

### Role of the funding source

The funders of the study had no role in the study design, data collection, data analysis, data interpretation, or writing of the report.

## RESULTS

After de-duplication a total of 10477 titles and abstracts were screened of which 8829 were deemed not relevant (figure 2). 145/1648 (8.8%) full texts could not be sourced. A further 1280 studies were deemed to meet exclusion criteria, leaving 223 for bias assessment. A high risk of bias was found in 109 studies and an additional 90 could not reliably have data extracted and therefore did not contribute to our results. In total, 22 English and two German articles, with a combined sample of 139,212 participants contributed 34 cohorts for analysis. Eight of the 24 studies scored maximal scores on the General Quality Assessment. The quality of data on symptom status was generally poor, with 10 studies scoring zero stars in the Specific Quality Assessment (supplementary table 2).

**Figure 2:**
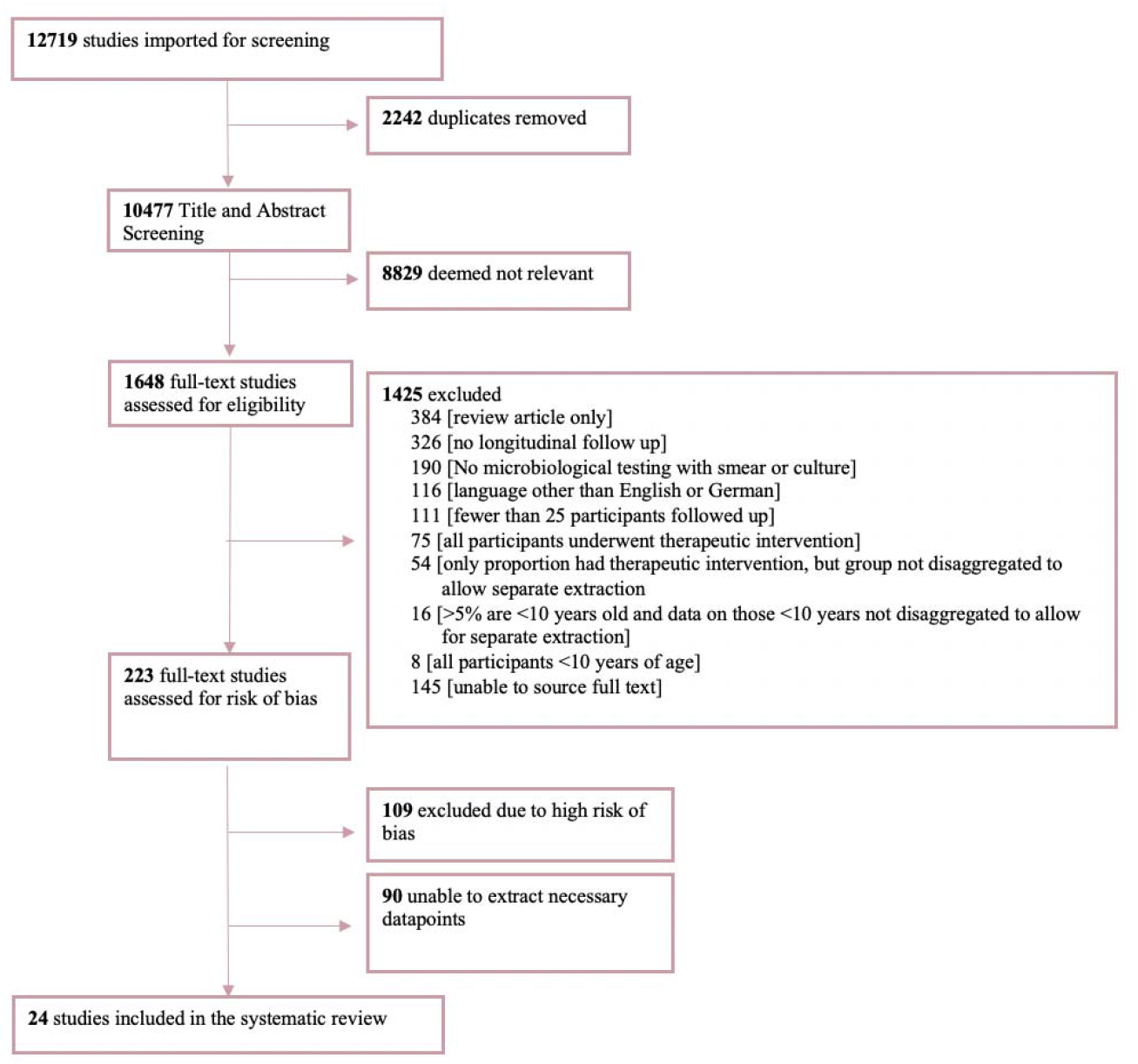
Study Selection.

The setting for the 34 longitudinal cohorts were as follows: workplace or university screening (n=5), general community screening (n=7), from household contact studies (n=4), clinical cohorts at clinics or sanatoria (n=9) and control arms of therapeutic interventions (n=9) (table 1 and supplementary table 3). Cohorts were conducted in Europe (n=10), Asia (n=11), North America (n=11), Africa (n=1) and South America (n=1). Eleven of the 34 cohorts provided an estimate of the local burden of TB disease in the study setting and related time period. The majority (n=9/11) of these settings would be classified as endemic or high burden TB settings, and the remainder (n=2/11) as medium burden, based on today’s WHO classification(19). Cohorts were conducted between 1923 and 2004 with 20/34 (58.8%) prior to 1960.

**Table 1:**
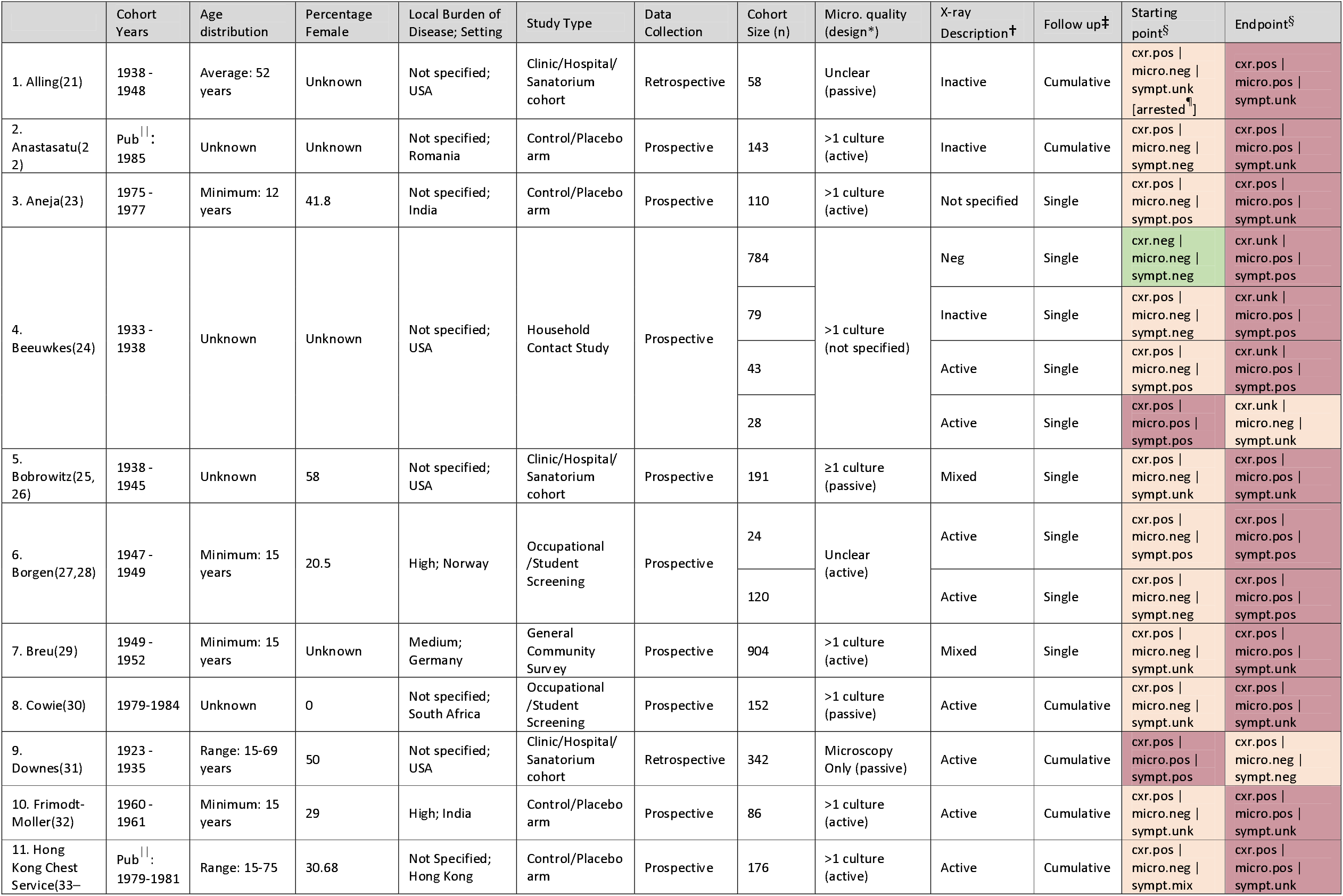
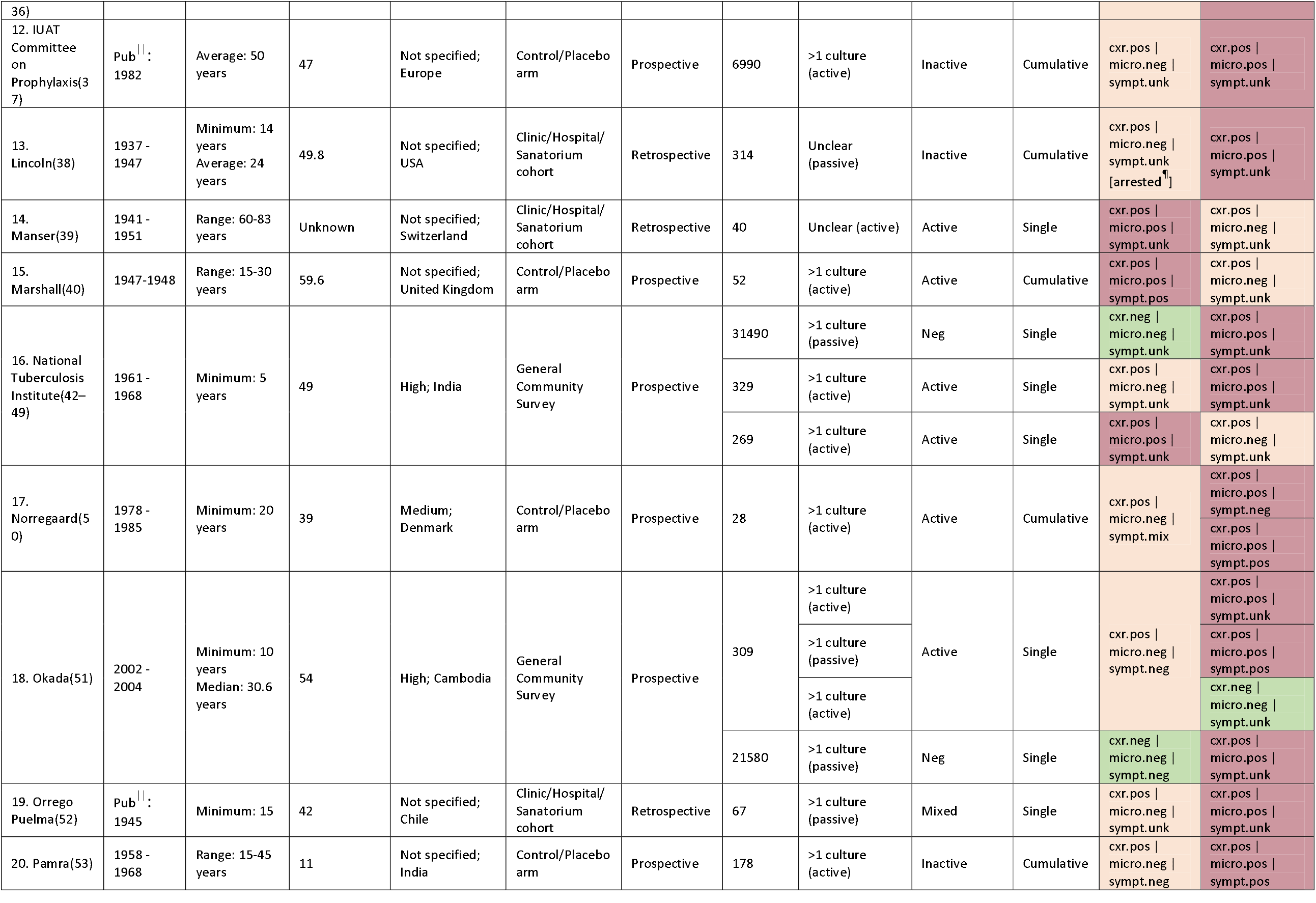
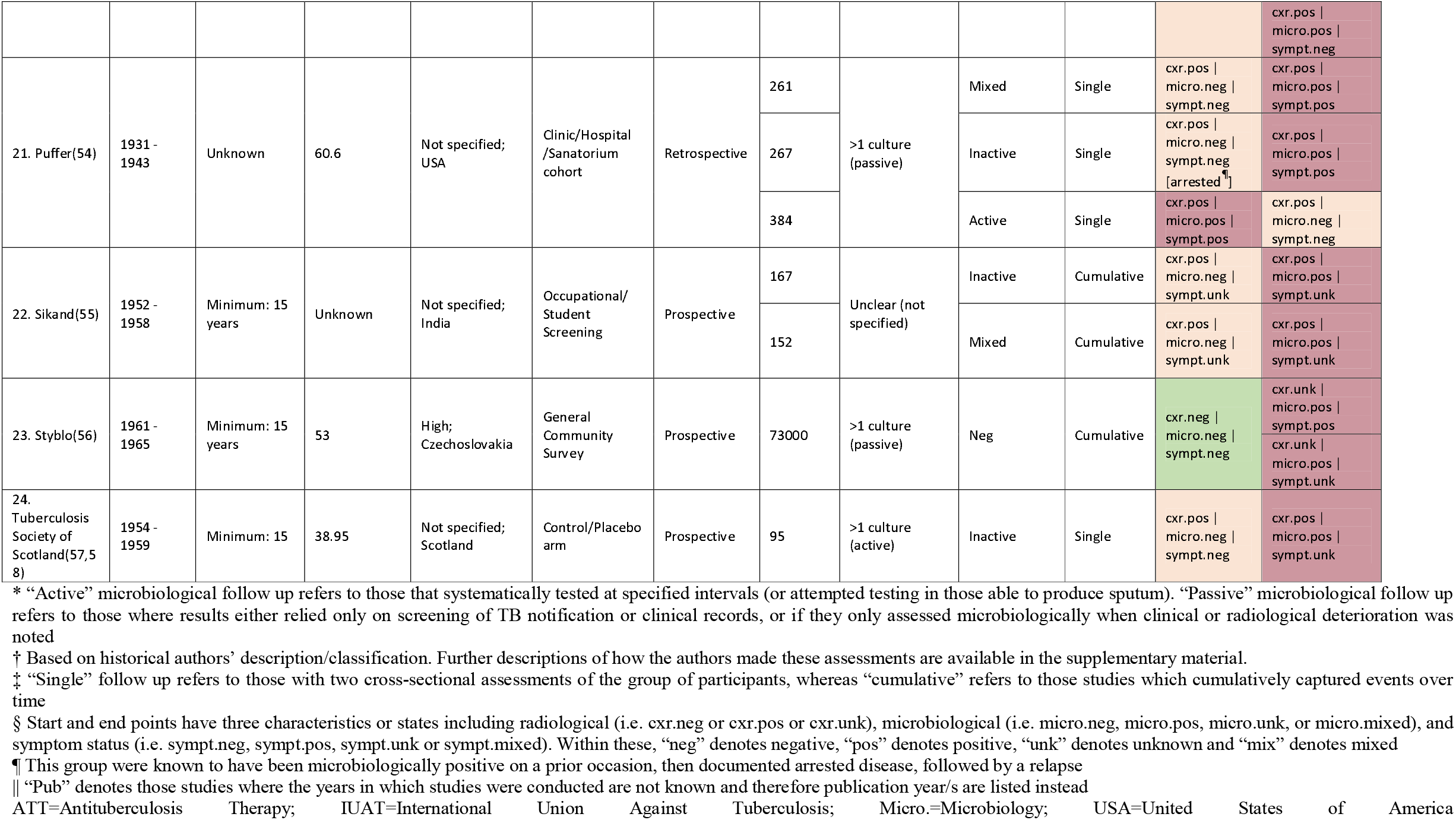
Extracted studies

We did not identify any cohorts, meeting our inclusion and quality criteria, closely following up confirmed recent TST converters where transition from normal chest X-ray (CXR) to CXR suggestive of TB was reported. We identified four cohorts following up participants with normal radiography, negative microbiological testing where the timepoint of initial infection was unclear, with either no evidence of symptoms (n=3 (75.0%)) or unrecorded symptom status (n=1 (25.0%)) (table 1). We identified 24 cohorts following-up participants with evidence of radiographic abnormalities and negative microbiology but with either no symptoms (n=8 (33.3%)), symptoms (n=3 (12.5%)) or mixed/unknown symptoms (n=13 (54.2%)) initially. Of these 24 cohorts, the radiographic abnormalities were specified by the original authors as either active (n=9 (37.5%)) or inactive/fibrotic (n=7 (33.3%)), with the remaining being mixed or not specified (n=8 (29.1%)). We identified six cohorts following participants with microbiologically detectable tuberculosis either initially with symptoms (n=4 (66.7%)) or those with an unknown symptom status (n=2 (33.3%)), however there were no cohorts found in which patients were documented to be asymptomatic.

### Progression to microbiologically positive disease in those with abnormal chest x-ray at baseline

From the 24 cohorts with abnormal chest radiography but no evidence of *M. tb* on respiratory sampling at baseline representing 11,185 participants, development of microbiologically-detectable disease occurred in between 1.1 – 57.9% of individuals with half the studies reporting a follow-up of up to three years (range 12-156 months) (figure 3a). Considerable statistical heterogeneity was seen across cohorts (I^2^ = 97.3%, tau^2^=0.001, p<0.01). We considered that the radiographic abnormalities categorized as active versus inactive TB (as specified by the original authors; supplementary table 4) could represent distinct pathological states contributing to clinical variability of studies. Therefore we did not pool these studies in meta-analysis, but rather conducted stratified meta-analysis to describe the progression of these two states separately. The annualized rate of transition from microbiologically negative to positive was 9.7% (95% CI: 6.2-13.3) for those in the nine cohorts described to have active changes on radiography compared to 1.1% (95% CI: 0.3-1.8) for those in the seven cohorts with inactive changes (figure 3a). Over a three-year period, this would equate to 26% (95%CI: 17-35) of those with active TB changes vs 3% (95%CI: 1-5) with inactive TB changes progressing from microbiologically positive to negative disease. Statistical heterogeneity in the active and inactive TB subgroups was lower than in all cohorts taken together, I^2^ = 77.4% and I^2^ = 53.2% respectively. The annual progression in cohorts with “mixed” radiographic changes was 6.3% (95% CI: 1.5-11.1) - in between the values for inactive and active strata.

**Figure 3a:**
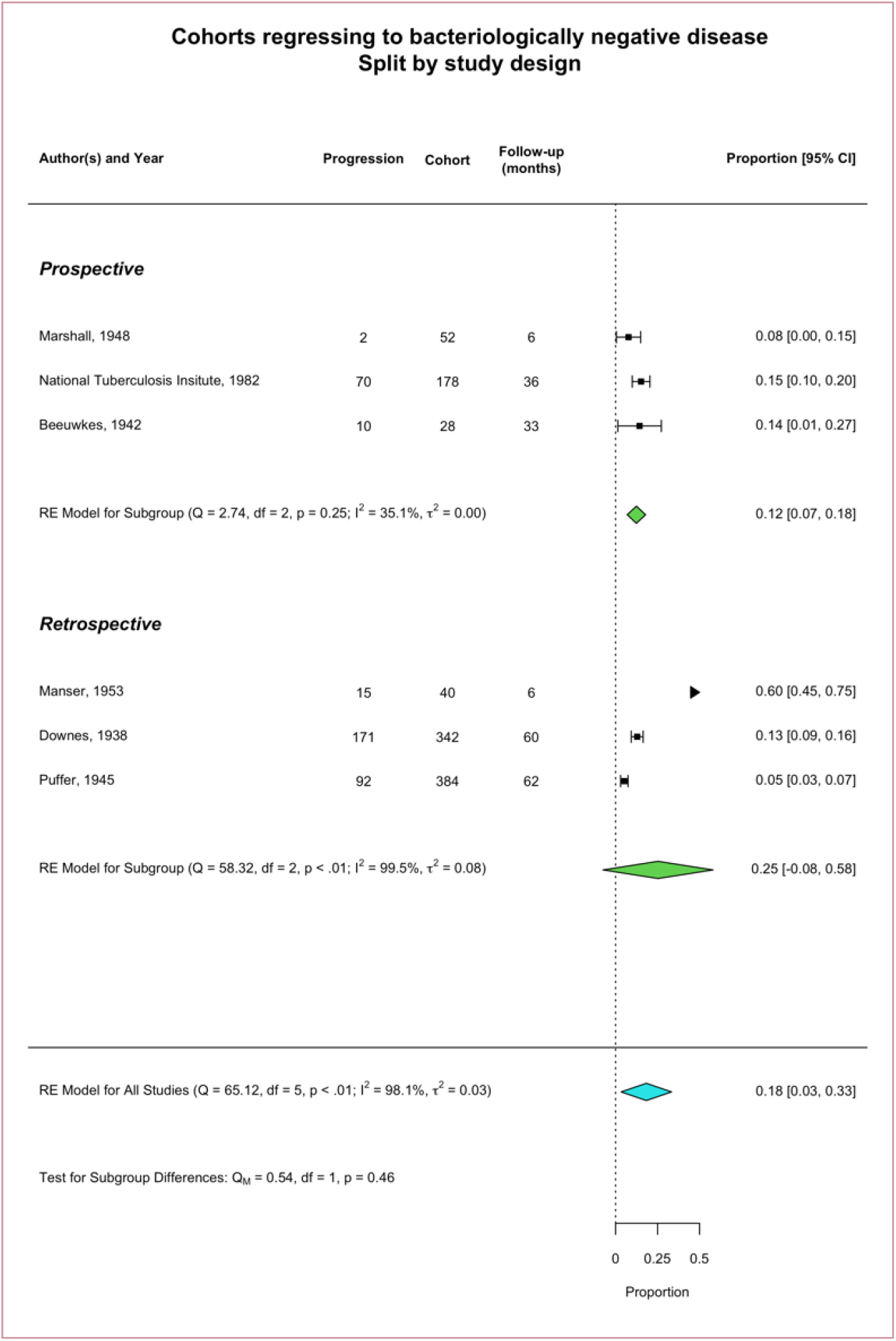
participants entering cohorts with abnormal chest X-rays and negative microbiology, transitioning to positive microbiology: forest plot of the random effects meta-analysis of annualized rates (as described fully in methods section) with proportion and 95% confidence intervals for subgroups. Subgroups are as per the historical authors’ provided data on radiographic classification being either “active”, “inactive” or where the group was “mixed”.

Out of 24 cohorts that contributed patients to this group, 18 (75%) used culture as part of microbiological work-up and the remainder (n=6/25) did not specify the microbiological tests undertaken. Restricting this analysis to the 18/24 cohorts explicitly using culture had little impact on these results (supplementary figure 1). Only 11 cohorts provided data on symptom status. Of the 9 cohorts described to have active TB changes on radiography, three were in symptomatic individuals. Progression in this subgroup was at an annualized rate of 11.74% (95% CI: 2.73-20.75) (supplementary figure 2). There was only one cohort describing active TB changes on radiography in an asymptomatic group with the remainder unknown.

In the four cohorts following up those with no radiographic changes suggestive of any TB (table 1), transition to microbiologically positive occurred at an annualized rate of 0.14% (95% CI: 0.11-0.17) (supplementary figure 3a). In “single follow up” and “cumulative count” studies, those with active TB changes showed similar annual progression (supplementary figure 4).

**Figure 3b:**
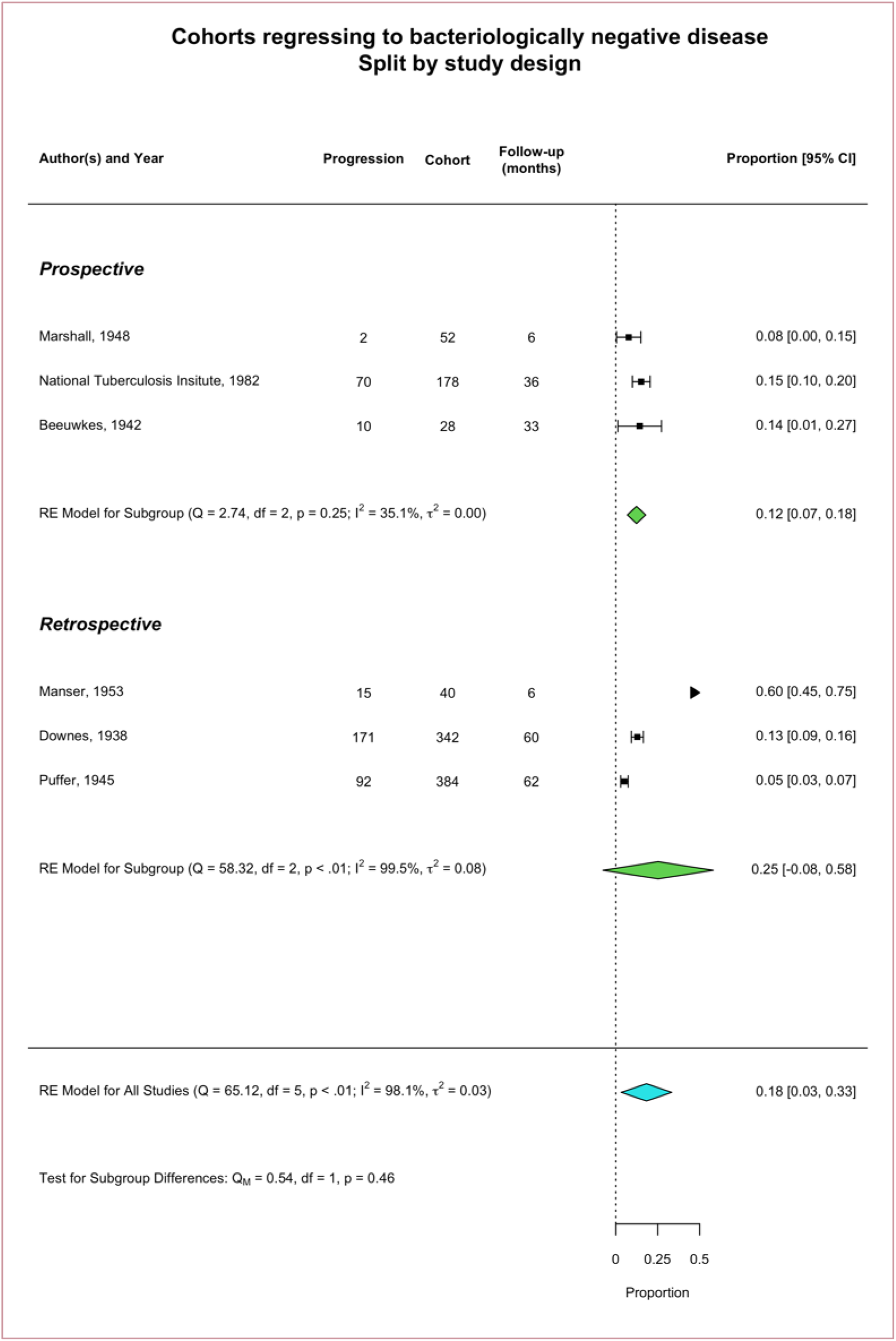
participants entering cohorts with positive microbiology, transitioning to negative microbiology: forest plot of the random effects meta-analysis of annualized rates (as described fully in methods section) with proportion and 95% confidence intervals for subgroups, according to study design

### Regression to negative microbiology in those with positive microbiology at baseline

Six cohorts followed a total of 1115 participants with evidence of *M. tb* in respiratory samples at baseline and assessed the proportion transitioning to a microbiologically undetectable state without treatment or intervention, with two thirds of studies reporting up to three years of follow up (range 6-26 months). The majority of these cohorts included participants with limited or minimal disease on CXR - either due to this being entry criteria into the original study or due to the eligibility criteria of this systematic review. No studies were able to adequately describe symptom status of the participants. Three out of six were retrospective cohorts from TB hospitals or sanatoria and three were prospective cohorts from general community/household surveys or a placebo arm of a trial. In four of the six cohorts, culture was used to assess microbiological status of participants while in two cohorts, both retrospective, either microscopy was used or nature of microbiological investigations was not specified. With meta-analysis, this transition occurred at an annualized rate of 18.3% (95% CI: 3.0-33.7) (figure 3b), but there was considerable heterogeneity across these studies (I^2^ = 98.1%, tau^2^=0.03, p<0.01). We then restricted the meta-analysis to prospective studies, hence removing the three retrospective hospital/sanitoria cohorts, where culture had also not be used in two instances, and showed an annualized rate of 12.4% (95% CI: 6.8-18.0) with reduced statistical heterogeneity I^2^ = 35.1%. Over three years this would equate to 33% (95% CI: 19-45) of those initially with culture positive TB becoming culture negative.

## DISCUSSION

This review is the first to systematically summarize key aspects of the kinetics of the natural history of untreated tuberculosis in adults, outside of the rate of mortality, making full use of historical literature in English and German. Through meta-analysis we provide estimates of the risk of progression to microbiologically positive disease in those with initially negative microbiology at an annualized rate of approximately 10% in those with “active” radiographic TB changes and 1% in those with “inactive” or fibrotic changes. For comparison, progression was approximately 0.1% for those with normal CXRs, while recognizing that this rate would be affected factors such as local burden of disease. In addition to this we provided an estimate for the reversion from culture-positive disease to culture negative without treatment (also referred to as ‘self-cure) as 12.4% per year.

These results highlight that individuals with CXR changes suggestive of active TB but who are found to be initially microbiologically negative are at considerable risk of disease progression. Guidance on how to manage this group is often unsatisfactory with a tension between providing empirical treatment, or monitoring for bacteriological progression which can be resource intensive or unfeasible in resource constrained settings. Better understanding of the risk of disease progression provided by this study could assist decision making and inform the planning of necessary clinical trials. In addition, our study is the first to determine an estimate for this transition which will be of use to modellers wanting to understand the implications on intervening in this population. We also have shown that approximately a third of those with culture positive disease could revert to culture negative without treatment over a 3-year period. While this may not inform clinical management, our results may refine parameters in models used to estimate disease incidence from prevalence survey data where the probability of so-called “self-cure” needs to be factored in. Our annual rate of approximately 12% provides empirical foundation to the slightly higher rates of 15% and 20% used by Dye to parameterize “self-cure” – which was informed by a review of literature although not systematic(59,60).

We used a widely accepted conceptual framework to guide our data collection which required determination of the microbiological, radiological and symptom status of participants over follow-up. We found that no single study systematically recorded these three features over the entire course of disease from exposure to final outcome. In addition we found that the recording of symptoms in these studies was not explicit, particularly during follow up - meaning there was insufficient empirical data to directly determine the trajectory around subclinical (asymptomatic, microbiologically positive) TB. Subclinical TB is a commonly identified state through CXR-based active case finding but conducting contemporary natural history studies to determine the rates of progression and regression would present ethical challenges with the availability of treatment. However, the substantial additional data uncovered in this review should allow inference of the kinetics around subclinical TB, which Richards *et al* have explored using a Bayesian framework to utilize the information from all available data simultaneously(61).

There are several key limitations to consider when interpreting the findings of this systematic review. HIV is a significant role-player in the epidemiology of TB in certain settings today and 22 of 24 of our studies were set prior to the discovery of the virus. It is likely that people living with HIV progress along the disease spectrum with different kinetics, also influenced by immune status (62–64). Secondly the nature of this research question and the historical focus resulted in studies being included from a period spanning almost 80 years; over this time period, microbiological and radiographic methods evolved. However, from a microbiological perspective included studies predominately used culture and where they did not, we conducted sensitivity analyses. For radiology, even where studies used mass miniature radiography or fluoroscopy for screening, findings were typically confirmed with conventional chest radiography which informed data extraction. The majority of studies were conducted over fifty years ago, when socioeconomic, health access, comorbidity distribution and prevalence of TB were likely very different to what they are today. However, these study environments may to a certain extent remain representative of many contemporary settings with a high TB burden.

There are also considerable methodological challenges in conducting a systematic review involving historical research. It is notable that 1503/1648 (91.2%) of studies were retrieved for full text review, however for 95 studies that met eligibility and bias criteria, manuscript style did not allow for data extraction and authors could not be contacted for assistance. Although our work focused on the period 1895-1960, through extensive investigator collections and snowballing of references we are confident we were able to identify key literature post-1960 as evidenced by nearly half of our final 24 studies being after this date.

Through our extensive review, we find that the natural history of TB is a dynamic, heterogenous process which is not adequately represented by a single ‘active disease’ state, and quantified three key transitions. Importantly, this review provides a much-needed foundation of empirical data for our ongoing re-discovery of the complexity of TB natural history, enabling a grounding for new preconceptions or dogmas, and a drive toward new clinical guidelines and policies for those suffering from TB.

## Data Availability

All data produced in the present work are contained in the manuscript

## CONTRIBUTORS

HE, RH, BS, AR, FC, and KK conceptualised the study protocol. BS, AR, TH, BF, FB, AO, and BH carried out the literature search and data collection. AR and BS carried out the statistical analysis and verified the final data with input and oversight from HE, RH and ER. BS wrote the first draft of the manuscript with input from AR, RH and HE. All authors subsequently reviewed and edited the manuscript. All authors had full access to the study data and had final responsibility for the decision to submit for publication.

## DECLARATION OF INTERESTS

We declare no competing interests.

## ACKNOWLEDGEMENTS

This work was funded by TB Modelling and Analysis Consortium (MAC), Medical Research Council (Grant ref: MR/V00476X/1) and the European Research Council (Starting Grant Action Number 757699). We would like to acknowledge Adrienne Burrough, Bridget Chivers and Rohisha Luchin for their assistance in performing the hand searches of Index Medicus and in collecting literature from various English libraries, and would like to thank the team of the German Tuberculosis Archives in Heidelberg for assisting with collection of German literature.

## Notes

### Competing Interest Statement

The authors have declared no competing interest.

### Clinical Protocols

https://www.crd.york.ac.uk/prospero/display_record.php?RecordID=152585

### Funding Statement

This study was funded by Bill and Melinda Gates Foundation via a grant to the TB Modelling and Analysis Consortium

