## Supplementary Materials for "The Natural History of Untreated Pulmonary Tuberculosis in Adults: A Systematic Review and Meta-Analysis"

### **Supplementary Appendix for:**
### Natural History of Untreated Pulmonary Tuberculosis in Adults: A Systematic Review and Meta-Analysis

Bianca Sossen (MBChB)[1,2,#], Alexandra Richards (MMathPhys)[3,4,#], Torben Heinsohn (BMBCh)[2,5], Beatrice Frascella (MD)[6], Federica Balzarini (MD)[6], Aurea Oradini-Alacreu (MD)[6], Prof. Anna Odone (PhD)[7], Ewelina Rogozinska (PhD)[8], Brit Häcker (Dr)[9], Prof. Frank Cobelens (PhD)[10,11], Prof. Katharina Kranzer (PhD)[12-14], Prof. Rein MGJ Houben (PhD)[3,4;&], Hanif Esmail (PhD)[2,8,15;&,¥]

[1] Department of Medicine, Faculty of Health Sciences, University of Cape Town, South Africa
[2] Institute for Global Health, University College London, London, UK
[3] TB Modelling Group, TB Centre, London School of Hygiene and Tropical Medicine, London, UK
[4] Department of Infectious Disease Epidemiology, Faculty of Epidemiology and Public Health, London School of Hygiene and Tropical Medicine, London, UK
[5] Helmholtz Centre for Infection Research, Department of Epidemiology
[6] School of Public Health, Vita-Salute San Raffaele University, Milan, Italy
[7] Department of Public Health, Experimental and Forensic Medicine, University of Pavia, Pavia, Italy
[8] MRC Clinical Trials Unit at University College London, UK
[9] German Central Committee against Tuberculosis (DZK), Berlin, German
[10] Amsterdam University Medical Centers location University of Amsterdam, Department of Global Health
[11] Amsterdam Institute for Global Health and Development, Amsterdam, The Netherlands
[12] Clinical Research Department, Faculty of Infectious and Tropical Disease, London School of Hygiene and Tropical Medicine, London, UK
[13] Biomedical Research and Training Institute, Harare, Zimbabwe
[14] Division of Infectious Diseases and Tropical Medicine, University Hospital, LMU Munich, Munich, Germany
[15] Wellcome Centre for Infectious Diseases Research in Africa, Institute of Infectious Diseases and Molecular Medicine, University of Cape Town, South Africa

### Contributed Equally
& Contributed Equally

#### Supplementary Table 1: Adapted Newcastle Ottawa Quality Assessment Tool

| GENERAL QUALITY ASSESSMENT | MAXIMUM STARS |
|---|---|
| **STUDY SELECTION** | |
| 1. Representativeness of 'exposed' cohort | |
| Truly representative of general population e.g. identified through prospective recruitment following mass screening or household screening | 1 star |
| Somewhat representative of general population e.g. identified through occupational screening | |
| Selected group e.g. sanatorium or hospital patients | 0 stars |
| No description of the derivation of the cohort | |
| 2. Selection of non-exposed cohort | |
| *NOT APPLICABLE* | |
| 3. Ascertainment of exposure* | |
| Documented TST conversion | 1 star |
| Exposure appropriately inferred from significant exposure to active TB e.g. in household or occupational setting | |
| No clear significant exposure to active TB | 0 stars |
| No description | |
| 4. Demonstration that outcome of interest not present at start of study* | |
| Yes - evidence of TB excluded by CXR at beginning of follow-up, or documented as not being present on CXR within previous 12 months | 1 star |
| Not stated | 0 stars |
| **COMPARABILITY** | |
| *NOT APPLICABLE* | |
| **OUTCOME** | |
| 5. Assessment of outcome | |
| Disease defined on the basis of microbiological demonstration of organism | 1 star |
| Disease defined without microbiological evidence | 0 stars |
| No description | |
| 6. Could intervention prevent development of outcome | |
| No medical or surgical treatment provided | 1 star |
| Medical or surgical treatment provided to a subgroup of clearly identified individuals analysed separately and no selection bias | |
| Medical or surgical treatment provided to a subgroup (<10%) and that treatment deemed unlikely to influence outcome | |
| Medical or surgical treatment provided to a large number of individuals OR unable to be analysed separately OR selection bias | 0 stars |
| 7. Was total follow-up long enough for outcomes to occur | |
| Yes - follow-up equal to or longer than 12 months | 1 star |
| No | 0 stars |
| 8. Adequacy of follow-up of cohorts | |
| Complete follow-up | 1 star |
| Participants lost to follow up unlikely to introduce bias: <20% of participants lost to follow-up per annum, or description provided for those lost, making introduction of bias unlikely | |
| Lost to follow-up rate > 20% per annum and no description of those lost | 0 stars |
| No statement | |

* These sections were only applicable to those cohorts of patients that started with CXR and microbiology negative for TB

| **ADDITIONAL SPECIFIC QUALITY ASSESSMENT**<br>*- If study meets general quality criteria above, used to determine if microbiological, radiographic and symptoms are recorded to sufficient quality* | MAXIMUM STARS |
|---|---|
| **RADIOGRAPHY** | |
| 1. Technology | |
| Radiography (X-Ray, Roentgenography) | 1 star |
| Mass Miniature radiography (Photofluorography, Aburography) | |
| Other (please specify) | 0 stars |
| 2. Diagnostic criteria used | |
| Clear use of recognised diagnostic criteria (e.g. ATS) | 1 star |
| Clear description of study specific methodology (not recognised diagnostic criteria) | |
| No or limited description of criteria or methodology | 0 stars |
| No imaging | |
| 3. Methodology | |
| Double or triple independent read | 1 star |
| Single reader | 0 stars |
| Not specified | |
| 4. Follow-up | |
| Complete follow-up | 1 star |
| <20% of participants per annum in follow-up did not receive imaging OR description for those not receiving imaging, making introduction of bias unlikely | |
| <80% followed up received imaging and no description of those lost | 0 stars |
| No statement | |
| **MICROBIOLOGY** | |
| 1. Technology | |
| Culture - solid | 1 star |
| Culture - liquid | |
| Smear - with concentration | |
| Smear - without concentration | |
| Guinea pig inoculation | |
| Other (please specify) | |
| Not specified | 0 stars |
| 2. Sampling method | |
| 24-hour sputum collection | 1 star |
| 48-hour sputum collection | |
| 72-hour sputum collection | |
| Induced sputum (e.g. physiotherapy or saline) | |
| Spot sputum collection (including <24 hour collection | |
| Gastric aspirate | |
| Nasal/Laryngeal/Tracheal swab | |
| Other (please specify) | |
| Not specified | 0 stars |
| 3. Follow-up | |

| | |
|---|---|
| Complete follow-up of those able to produce sputum | 1 star |
| <20% of per annum of participants had sputum collected (of those able to produce) for microbiological investigation OR description provided for that lost, making introduction of bias unlikely | |
| Follow-up rate <80% and no description of those lost | 0 stars |
| No statement | |
| **SYMPTOMS** | |
| 1. Methodology | |
| Standardised symptom screen described in methodology | 1 star |
| Clear statement of symptomatic vs asymptomatic | |
| Standardised definitions from published criteria used with symptoms included in definition | |
| No clear statement | 0 stars |
| 2. Follow-up | |
| Complete follow-up | 1 star |
| <20% of participants per annum in follow-up did not get symptom assessment OR description provided for those lost, making introduction of bias unlikely | |
| Follow-up rate <80% and no description of those lost | 0 stars |
| No statement | |

### Supplementary Table 2: Bias Assessment

| | General Quality Assessment† | | Specific Quality Assessment | | |
|---|---|---|---|---|---|
| | Study Selection‡ | Outcome Assessment | Radiography | Microbiology | Symptoms |
| 1. Alling(21) | 0 | 0 0 0 X | 0 0 0 X | 0 0 X | 0 X |
| 2. Anastasatu(22) | X | 0 0 0 0 | 0 0 X X | 0 0 0 | X X |
| 3. Aneja(23) | 0 | 0 0 0 0 | 0 0 0 0 | 0 0 0 | 0 0 |
| 4. Beeuwkes(24) | 0 | 0 0 0 X | 0 0 X X | 0 0 X | 0 X |
| 5. Bobrowitz(25,26) | X | 0 0 0 0 | 0 0 0 0 | 0 0 0 | 0 0 |
| 6. Borgen(27,28) | 0 0 0 | 0 0 0 0 | 0 0 0 X | 0 0 X | 0 0 |
| 7. Breu(29) | 0 | 0 0 0 0 | 0 0 0 X | 0 0 0 | X X |
| 8. Cowie(30) | 0 | 0 0 0 0 | 0 0 0 X | 0 0 0 | X X |
| 9. Downes(31) | X | 0 0 0 X | 0 0 0 X | X X X | 0 0 |
| 10. Frimodt-Moller(32) | 0 | 0 0 0 0 | 0 0 0 0 | 0 0 0 | X X |
| 11. Hong Kong Chest Service(33–36) | X | 0 0 0 0 | 0 0 0 X | 0 0 0 | 0 0 |
| 12. IUAT Committee on Prophylaxis(37) | X | 0 0 0 0 | 0 0 0 X | 0 0 0 | X X |
| 13. Lincoln(21,38) | 0 | 0 0 0 X | 0 0 0 X | 0 0 X | 0 X |
| 14. Manser(39) | X | 0 0 0 X | 0 0 0 X | 0 0 0 | 0 0 |
| 15. Marshall(40) | 0 | 0 0 0 0 | 0 0 0 0 | 0 0 0 | 0 0 |
| 16. National Tuberculosis Institute(41–49) | 0 0 0 | 0 0 0 0 | 0 0 0 0 | 0 0 0 | X X |
| 17. Norregaard(50) | X | 0 0 0 0 | 0 0 0 X | 0 0 0 | 0 X |
| 18. Okada(51) | 0 0 X | 0 0 0 0 | 0 0 0 0 | 0 0 0 | 0 0 |
| 19. Orrego Puelma(52) | X | 0 0 0 0 | 0 0 0 X | 0 0 0 | 0 X |
| 20. Pamra(53) | 0 | 0 0 0 0 | 0 0 0 X | 0 0 0 | X X |
| 21. Puffer(54) | 0 | 0 0 0 X | 0 0 0 X | X X X | X X |
| 22. Sikand(55) | 0 0 0 | 0 0 0 X | 0 0 0 X | 0 0 0 | X X |
| 23. Styblo(56) | 0 0 0 | 0 0 X X | 0 0 0 X | 0 0 X | 0 X |
| 24. Tuberculosis Society of Scotland(57,58) | X | 0 0 0 0 | 0 0 0 X | 0 0 0 | X X |

* This table only includes the quality assessments for studies included in the meta-analysis, with a 0 representing a positive score and X representing a negative score.
† Studies could only lose two stars in the General Quality Assessment to proceed to data extraction. Further details of the quality assessment tools are available in supplementary material.
‡ Studies could score a maximum of 3 stars if participants entered the cohort with no evidence of disease (chest x-ray negative, microbiologically negative and asymptomatic) or a maximum of 1 star if participants already had some evidence of disease at entry.

#### Supplementary Table 3: Extracted studies

| | Population | Study Type | Dates of study | Imaging Assessment | Microbiological Assessment | Symptom Assessment | Follow up methods | Transition* | Proportion making transition (time, months) |
|---|---|---|---|---|---|---|---|---|---|
| **1. Alling(1)** (1955)<br><br>USA | • Diagnosis of moderate tuberculosis<br>• Followed up within the study hospital<br>• Sourced from referral from private physician or mass occupational radiographic surveys | Clinic/ Hospital/ Sanatorium Cohort | 1938 - 1948 | Chest X-ray (CXR) reviewed according to NTA† criteria with double, independent reading. All "moderate" tuberculosis at baseline | Sputum sent for unknown method. All deemed "arrested" in this data row. | Delineation of symptom status is not well defined but all individuals in this data row were deemed "arrested" | Retrospective collection of routinely collected clinical data from their hospital. Follow up was therefore not systematically done. Definitions of disease groups required repeat CXR and sputum testing over periods of time. | cxr.pos \| micro.neg \| sympt.unk [arrested‡]<br><br>to<br><br>cxr.pos \| micro.pos \| sympt.unk | • 8/58 (60)<br>• 10/58 (60-156) |
| **2. Anastasatu(2)** (1985)<br><br>Romania | Smear negative with minimal or moderately advanced pulmonary lesions | Control/ Placebo arm | Unknown | CXR | Sputum smear and culture | Not specified but described to be "without radio-clinical symptoms of activity" | Follow up details not clear but authors describe follow up to 24 months, with sputum testing being performed | cxr.pos \| micro.neg \| sympt.neg<br><br>to<br><br>cxr.pos \| micro.pos \| sympt.unk | 6/143 (24) |
| **3. Aneja(3)** (1979)<br><br>India | • Symptomatic for TB and abnormal photofluorogram<br>• Negative sputum microscopy<br>• No TB treatment, or less than 2 weeks of treatment prior to entry into trial<br>• Well enough for ambulant care | Control/ Placebo arm | 1968 - 1972 | CXR that was double-read ± a third umpire in case of discrepancies | Sputum for microscopy and culture | Review of clinical symptoms e.g. cough | Repeat CXR and spot sputum for microscopy and culture were done at the end of the 2nd, 4th, 6th, 9th and 12th months of follow up | cxr.pos \| micro.neg \| sympt.pos<br><br>to<br><br>cxr.pos \| micro.pos \| sympt.unk | 21/110 (12) |
| **4. Beeuwkes(4)** (1942) | Household contacts with at least 6 months of follow up data | Household Contact Study | 1933-1938 | CXR | 72 hour concentrated sputum sample for microscopy. Some also had | Based on medical history and examination to have been "asymptomatic" | Re-examined 6-12 monthly plus additional unscheduled visits if | cxr.neg \| micro.neg \|symp.neg<br><br>to | 1/784 (6-60) |

| | | | | | | | | | |
|---|---|---|---|---|---|---|---|---|---|
| USA | | | | | culture and/or inoculation. | | developing symptoms.<br>Reviews included medical history and examination, chest x-ray and sputum sampling. | cxr.unk \| micro.pos \| sympt.pos | |
| | | | | | | | | cxr.neg \| micro.neg \| symp.neg<br><br>to<br><br>cxr.unk \| micro.neg \| sympt.pos | 3/784 (6-60) |
| | | | | | | | | cxr.pos \| micro.neg \| sympt.neg<br><br>to<br><br>cxr.unk \| micro.pos \| sympt.pos | 3/79 (6-60) |
| | | | | | | | | cxr.pos \| micro.neg \| sympt.neg<br><br>to<br><br>cxr.unk \| micro.neg \| sympt.pos | 5/79 (6-60) |
| | | | | | | | | cxr.pos \|. micro.neg \| sympt.neg<br><br>to<br><br>cxr.unk \| micro.pos \| sympt.pos | 13/43 (6-60) |
| | | | | | | | | cxr.pos \| micro.pos \| sympt.pos<br><br>to<br><br>cxr.unk \| micro.neg \| sympt.unk | 10/28 (6-60) |
| **5. Bobrowitz(5, 6)** (1947; 1949)<br><br>USA | • Patients at study site with minimal disease on Chest X-ray<br>• At least 6 months of follow up data | Clinic/ Hospital/ Sanatorium Cohort | 1938 - 1945 | CXR | Sputum sample or gastric concentrates or cultures (authors noted that gastric cultures became more frequent | Symptoms and clinical review | Radiographic, mycobacteriological and clinical review during Sanatorium stay but intervals not clearly defined | cxr.pos \| micro.neg \| sympt.unk<br><br>to<br><br>cxr.pos \| micro.pos \| sympt.unk | 26/191 (60) |

| | | | | | | | | | |
|---|---|---|---|---|---|---|---|---|---|
| | | | | | from 1939 onwards) | | | | |
| 6. **B**o**rgen(7,8)** (1951;1952)<br><br>Norway | • Age: >15 years<br>• Factory workers who participated in a repeat survey | Occupation al/ Student Screening | 1947 - 1949 | Photofluorogra phy: no lesion versus suggestive tuberculous lesion | It is unclear whether systematic sputum testing was done at baseline, irrespective of photofluorograph y results | Clinical review including symptoms such as loss of weight and signs such as pyrexia | Factory workers had repeat survey at 2 years follow up point. Included repeat photofluorography. Also included sputum testing but unclear as to whether this was performed in all or only those with radiographic abnormalities. | cxr.neg \| micro.unk \| sympt.unk<br><br>to<br><br>cxr.pos \| micro.pos \| sympt.pos | 4/6884 (30) |
| | | | | | | | | cxr.pos \|. micro.neg \| sympt.pos<br><br>to<br><br>cxr.pos \| micro.pos \| sympt.pos | 2/24 (30) |
| | | | | | | | | cxr.pos \| micro.neg \| sympt.neg<br><br>to<br><br>cxr.pos \| micro.pos \| sympt.pos | 2/120 (30) |
| **7. Breu(9)** (1954)<br><br>Germany | • Patients that were assessed as part of mass screening in Ludwigsburg, Germany<br>• Demonstrated abnormal chest X-ray at baseline but micro negative | General Community Survey | 1949 - 1952 | Photofluorogra phy, CXR, tomography | Sputum for culture if either symptomatic or CXR abnormality suggestive of TB (but not all patients were able to produce sputum) | Delineation of symptom status is not well defined. All patients had an ESR performed at baseline. | Followed up "regularly" but interval not specified. Follow up included CXR, CT, microbiology (for sputum, laryngeal swab, gastric lavage) | cxr.pos \| micro.pos \| sympt.unk<br><br>to<br><br>cxr.pos \| micro.pos \| sympt.unk | 48/904 (3-48) |
| **8. Cowie(10)** (1985)<br><br>South Africa | • New or enlarging apical lesions on 6-monthly Chest X-ray occupational screening<br>• Apical lesion still present on repeat Chest X-ray 2 months later<br>• Concentrated smear X3 (baseline) and culture X2 (2-months later) of sputum was | Occupation al/ Student Screening | 1979 - 1984 | CXR with new or enlarging apical lesions | Early morning sputa for concentrated microscopy and culture | The authors do not comment on the clinical status/symptoms of the cohort | • 3-monthly CXRs for 3 years followed by 6-monthly until 5 year endpoint in the study<br>• Sputum for smear or culture if there was any progression radiographically or if the original lesion was thought to suggest bacteriologically positive disease<br>• Histology from | cxr.pos \| micro.neg \| sympt.unk<br><br>to<br><br>cxr.pos \| micro.pos \| sympt.unk | 88/152 (3-58) |

| | | | | | | | | | |
|---|---|---|---|---|---|---|---|---|---|
| | negative for MTB<br>• Employee at the Gold Mine | | | | | | bronchoscopy was also performed in some | | |
| **9. Downes(11)** (1938)<br><br>USA | • Resident of Cattaraugus County<br>• Diagnosed with active TB at some stage during the cohort years | Clinic/ Hospital/ Sanatorium Cohort | 1923 - 1935 | CXR | Microscopy on sputum (to confirm certain definitions, sputum was also concentrated) | Met the criteria for 'active' disease which was based on symptoms and examination | Retrospective data collection of clinical records, sanatoria outcomes and self-reporting. Data collection included that of mycobacteriology, clinical status and imaging. | cxr.pos \| micro.pos \| sympt.pos<br><br>to<br><br>cxr.pos \| micro.neg \| sympt.neg | • 27/342 (12)<br>• 104/342 (24)<br>• 140/342 (36)<br>• 158/342 (48)<br>• 171/342 (60) |
| **10. Frimodt-Moller(12)** (1965)<br><br>India | • Abnormalities on Chest X-ray at baseline (and deemed probably tuberculous aetioloygy by two reviewers)<br>• Negative smear and culture from sputum and larryngeal swab | Control/ Placebo arm | 1960-1961 | CXR deemed probably tuberculous by two readers | Microscopy and culture of sputum and laryngeal swab | Delineation of symptom status is not well-defined | 3-monthly follow-ups occurred over a 3 year period including CXR and microbiological assessment | cxr.pos \| micro.neg \| sympt.unk<br><br>to<br><br>cxr.pos \| micro.pos \| sympt.unk | • 11/86 (12)<br>• 18/86 (24)<br>• 25/86 (36) |
| **11. Hong Kong Chest Service**(13–16)<br>(1979, 1981, 1981, 1984)<br><br>HONG KONG | • Age: 15-75 years<br>• Radiographic evidence of 'active pulmonary tuberculosis' by a Hong Kong Chest Physician (not those considered fibrotic/inactive)<br>• No previous ATT<br>• At least 5 sputum smears negative over approximately one week | Control/ Placebo arm | Not specified | CXR | Microscopy and culture of sputum X 5 | Cough, sputum production, haemoptysis | • 1-2 sputum specimens for microscopy and culture at monthly reviews in the first year and then quarterly reviews up to 30 months<br>• 20 CXRs done between baseline and month 60<br>• Clinical reports made at each sampling/imaging visit<br>• Unscheduled additional reviews | cxr.pos \| micro.neg \| sympt.mixed<br><br>to<br><br>cxr.pos \| micro.pos \| sympt.unk | • 40/176 (3)<br>• 49/176 (6)<br>• 61/176 (12)<br>• 67/176 (18)<br>• 69/176 (24)<br>• 70/176 (30)<br>• 71/176 (36)<br>• 71/176 (60) |

| | | | | | | | | | |
|---|---|---|---|---|---|---|---|---|---|
| | | | | | | | with imaging & sputum sampling occurred if deterioration suspected | | |
| **12. IUAT Committee on Prophylaxis(17)** (1982)<br><br>Europe | • Fibrotic lesions on Chest X-ray that had been stable during the year prior to study entry<br>• Positive Mantoux<br>• No previous ATT<br>• Not previously mycobacteriologically positive and not culture positive at entry to the trial | Control/ Placebo arm | Unknown | CXR showing fibrotic lesions | 2 Sputum cultures at entry | Not well-defined | • Annual reviews were done for 5 years including sputum culture, CXR and/or vital status was recorded<br>• Records were also reviewed annually to see whether participants had been started on TB treatment | cxr.pos \| micro.neg \| sympt.unk<br><br>to<br><br>cxr.pos \| micro.pos \| sympt.unk | 97/6990 (12-60) |
| **13. Lincoln(18)** (1954)<br><br>USA | • No history of prior diagnosis of TB<br>• Now Chest X-ray suggestive of minimal TB<br>• Followed up within the study hospital for at least one year<br>• Sourced from referral from private physician (due to symptoms or a known TB contact) or from mass occupational radiographic surveys | Clinic/ Hospital/ Sanatorium Cohort | 1937 - 1947 | CXR reviewed according to NTA criteria with double, independent reading. All "minimal" tuberculosis at baseline in this data row. | Sputum sent for unknown method. 128/134 had sputum examination for tubercle bacilli. 112/128 had at least one "positive" sputum by some method. | The authors' definition of 'active' disease makes use of the NTA definitions which include symptom activity within their definitions. The majority of those with 'active' disease were also noted to have self-presented for care rather than having being picked up during general community screening. | Retrospective collection of routinely collected clinical data from their hospital. Follow up was therefore not systematically done. Definitions of disease groups required repeat CXR and sputum testing over periods of time. | cxr.pos \| micro.mixed \| sympt.pos<br><br>to<br><br>cxr.pos \| micro.neg \| sympt.unk | • 45/134 (24)<br>• 71/134 (36)<br>• 80/134 (48)<br>• 83/134 (60)<br>• 86/134 (72)<br>• 87/134 (84) |
| | | | | | | | | cxr.pos \| micro.neg \| sympt.unk [arrested‡]<br><br>to<br><br>cxr.pos \| micro.pos \| sympt.unk | • 15/314 (24)<br>• 25/314 (36)<br>• 32/314 (48)<br>• 35/314 (60)<br>• 36/314 (72) |
| **14. Manser(19)** (1953)<br><br>Switzerland | • Patients from the study sanatorium's records<br>• Over 60 years of age | Clinic/ Hospital/ Sanatorium Cohort | 1941 - 1951 | CXR | Sputum or gastric lavage for unspecified method | Symptom questioning, fever, temperature, weight; but the authors do not | Frequency of follow up was not specified, but included physical exam, weight, ESR, CXR/CT, and sputum or gastric lavage | cxr.pos \| micro.pos \| sympt.unk<br><br>to | 15/40 (6) |

| | | | | | | | | | |
|---|---|---|---|---|---|---|---|---|---|
| | | | | | | present breakdown of data | | cxr.pos \| micro.neg \| sympt.unk | |
| **15. Marshall(20)** (1948)<br><br>UK | • Acute bilateral pulmonary tuberculosis of presumably recent origin<br>• Old-standing disease and disease with thick-walled cavities excluded<br>• Bacteriologically proven<br>• Unsuitable for collapse therapy<br>• 15 to 30 years of age | Control/ Placebo arm | 1947 - 1948 | CXR | Sputum (direct smear and culture) ± laryngeal swab or gastric lavage if unable to produce (for culture) | General condition, temperature, weight | • All remained admitted to the sanatorium for 6 months and outcomes were assessed at this point<br>• Follow up included monthly clinical reviews<br>• CXR (by blinded radiologist) and mycobacterial investigations done by clinical team were recorded | cxr.pos \| micro.pos \| sympt.pos<br><br>to<br><br>cxr.pos \| micro.neg \| sympt.unk | 1/52 (6) |
| **16. National Tuberculosis Institute**(21–28) (1974; 1976; 1976; 1976; 1978; 1978; 1978; 1978; 1982)<br><br>India | • Randomly selected households in Bangalore district entered into a mass survey<br>• Excluded those with BCG scars | General Community Survey | 1961 - 1968 | CXR (this data row: normal or "considerred to be of nontuberculous etiology") | Sputum (spot & overnight) for microscopy and culture in those with abnormal CXR or CXR not interpretable | The authors do not comment on the clinical status/symptoms of the cohort | Surveyed every 18-24 months including:<br>• Photofluorography<br>• Two sputum specimens if any history of abnormal CXR or CXR not interpretable | cxr.neg \| micro.neg \| sympt.unk<br><br>to<br><br>cxr.pos \| micro.pos \| sympt.unk | • 44/31490 (18)<br>• 99/17936 (60) |
| | | | | CXR (this data row: "suggestive of tuberculous etiology") | | | | cxr.pos \| micro.neg \| sympt.unk<br><br>to<br><br>cxr.pos \| micro.pos \| sympt.unk | • 23/329(18)<br>• 36/271 (60) |
| | | | | CXR | | | | cxr.pos \| micro.pos \| sympt.unk<br><br>to<br><br>cxr.pos \| micro.neg \| sympt.unk | • 86/269 (18)<br>• 70/178 (36) |
| **17. Norregaard(29)** (1990)<br><br>Denmark | • Abnormal Chest X-ray<br>• Sputum smear-negative | Control/ Placebo arm | 1978 - 1985 | CXR | Sputum or gastric lavage for microscopy and culture X 6 | Medical history: e.g. cough, haemoptysis, fever | CXR and culture of sputum or gastric lavage monthly for the first three months, followed by every second month | cxr.pos \| micro.neg \| sympt.mixed<br><br>to<br><br>cxr.pos \| micro.pos \| sympt.neg | 6/28 (48) |

| | | | | | | | | | |
|---|---|---|---|---|---|---|---|---|---|
| | | | | | | | for six months and annually thereafter for at least three years. In this data row: "slight or no symptoms" were deemed asymptomatic. | cxr.pos \| micro.neg \| sympt.mixed<br><br>to<br><br>cxr.pos \| micro.pos \| sympt.pos | 2/28 (48) |
| **18. Okada(30)** (2012)<br><br>Cambodia | • >10 years of age<br>• Living in survey area<br>• Two culture-negative specimens | General Community Survey | 2002 - 2004 | CXR that was deemed "TB suggestive" | Sputum for smear and solid culture X 2 | Medical history and examination | Reviewed TB registers for the development of incident TB cases | cxr.pos \| micro.neg \| sympt.neg<br><br>to<br><br>cxr.pos \| micro.pos \| sympt.pos | 5/309 (1-24) |
| | | | | | | | Conducted a repeat review at 2 years of all those with abnormal CXR at baseline, including medical history, CXR and two sputum specimens for smear and culture | cxr.pos \| micro.neg \| sympt.neg<br><br>to<br><br>cxr.pos \| micro.pos \| sympt.unk | 46/309 (24) |
| | | | | CXR that was deemed to be normal (n=20407) or to have an abnormality that was not suggestive of TB (n=1173) | | | Reviewed TB registers for the development of incident TB cases | cxr.neg \| micro.neg \| sympt.neg<br><br>to<br><br>cxr.pos \| micro.pos \| sympt.unk | 32/21580 (1-24) |
| | | | | CXR that was deemed "TB suggestive" | | | Conducted a repeat review at 2 years of all those with abnormal CXR at baseline, including medical history, CXR and two sputum specimens for smear and culture | cxr.pos \| micro.neg \| sympt.neg<br><br>to<br><br>cxr.neg \| micro.neg \| sympt.unk | 26/309 (24) |
| **19. Orrego Puelma(31)** (1945) | • Abnormal CXR of minimal extent<br>• Had at least 2 years of follow up | Clinic/ Hospital/ Sanatorium cohort | Unknown | CXR | Sputum and/or gastric washings for microscopy, | Cough, loss of weight, haemoptysis, back pain | Repeat CXR and sputum testing were done at unspecified | cxr.pos \| micro.neg \| sympt.unk<br><br>to | 18/67 (24) |

<table>
<tr><td>CHILE</td><td>data available for retrospective review</td><td></td><td></td><td></td><td>culture and/or inoculation</td><td></td><td>intervals over the two years</td><td>cxr.pos | micro.pos | sympt.unk</td><td></td></tr>
<tr><td rowspan="2">20. Pamra(32) (1971)<br><br>India</td><td rowspan="2">• no history of previous treatment for Tb<br>• No manifest tuberculous lesion in any organ other than the lungs<br>• No evidence of diabetes or any other non-tuberculous disease<br>• Apparent radiological stability of the lesions during the period of observation before inclusion in the study, this period being not less than 3 months or more than 3 years<br>• Constantly negative results on direct smear and culture examination of at least 2 sputum and laryngeal swab specimens during the period of observation</td><td rowspan="2">Control/ Placebo arm</td><td rowspan="2">1958 - 1968</td><td rowspan="2">CXR</td><td rowspan="2">At least 2 sputum and laryngeal swab specimens smear and culture negative</td><td rowspan="2">Not well-described</td><td rowspan="2">• Laryngeal swab culture and CXR were done 3-monthly during year 1<br>• Laryngeal swab culture and CXR were done 6-monthly during year 2-5</td><td>cxr.pos | micro.neg | sympt.neg<br><br>to<br><br>cxr.pos | micro.pos | sympt.pos</td><td>2/178 (72)</td></tr>
<tr><td>cxr.pos | micro.neg | sympt.neg<br><br>to<br><br>cxr.pos | micro.pos | sympt.neg</td><td>55/178 (72)</td></tr>
<tr><td rowspan="2">21. Puffer(33) (1945)<br><br>USA</td><td rowspan="2">• Following up at study clinic at the time of study<br>• Deemed to have “reinfection type” Tuberculosis</td><td rowspan="2">Clinic/ Hospital/ Sanatorium cohort</td><td rowspan="2">1931 - 1943</td><td>CXR (in this data row, all deemed to have apical disease)</td><td rowspan="2">As per Opie criteria: "Arrested" or "Apparently arrested" is diagnosed when all symptoms have disappeared for a period of at least 6 months and the physical signs are those of</td><td rowspan="2">Review of “physical signs and/or symptoms”</td><td rowspan="2">Outcome for all study participants was assessed at 1 Jan 1943 irrespective of when they started in the study. It is not clear if follow up review was systematic or only based on routinely collected clinical data. In order to meet</td><td>cxr.pos | micro.neg | sympt.neg<br><br>to<br><br>cxr.pos | micro.pos | sympt.pos</td><td>9/261 (62)</td></tr>
<tr><td>CXR</td><td>cxr.pos | micro.neg | sympt.neg [arrested‡]<br><br>to</td><td>10/267 (62)</td></tr>
</table>

| | | | | | | | | | |
|---|---|---|---|---|---|---|---|---|---|
| | | | | | a healed lesion, the disease is regarded as arrested or, when some uncertainty still exists, the designation "apparently arrested" has been regarded as preferable. When the patient first comes under observation of the clinic, arrested lesions are not infrequently recognisable only by radiological examination, but are separable from "latent lesions" because they are known to have passed through a period of manifest disease with the usual symptoms of pulmonary tuberculosis. When, however, a tuberculous pulmonary lesion, recognised by roentgenographic examination, has been accompanied by no symptoms of the disease discoverable by inquiry, but is associated with physical signs of a | | study definitions, follow up would have had to meet NTA criteria. | cxr.pos \| micro.pos \| sympt.pos | |
| | | | | CXR | | | | cxr.pos \| micro.pos \| sympt.pos<br>to<br>cxr.pos \| micro.neg \| sympt.neg | 92/384 (62) |

<table>
<tr><td></td><td></td><td></td><td></td><td></td><td>healed lesion, such as impaired resonance or diminished breath sound, it should be classified as arrested tuberculosis"</td><td></td><td></td><td></td><td></td></tr>
<tr><td rowspan="3">22. Sikand(34) (1959)<br><br>India</td><td rowspan="3">Employed as police officer in study setting</td><td rowspan="3">Occupation al/ Student Screening</td><td rowspan="3">1952 - 1958</td><td>CXR deemed ‘inactive’ tuberculosis or tuberculosis of ‘doubtful activity’</td><td rowspan="3">Sputum or laryngeal swab for unknown method</td><td rowspan="3">Delineation of symptom status is not well defined</td><td rowspan="3">• CXR repeated at two timepoints:<br>o 17 months and<br>o 5-6 years<br>• Indications and regularity of mycobacteriological testing not clearly described</td><td>cxr.pos | micro.neg | sympt.unk<br><br>to<br><br>cxr.pos | micro.pos | sympt.unk</td><td>38/319 (12)</td></tr>
<tr><td rowspan="2">CXR deemed 'Negative' for TB</td><td>cxr.neg | micro.unk | sympt.unk<br><br>to<br><br>cxr.pos | micro.pos | sympt.unk</td><td>89/11268 (17-69)</td></tr>
<tr><td>cxr.neg | micro.unk | sympt.unk<br><br>to<br><br>cxr.pos | micro.neg | sympt.unk</td><td>251/11268 (17-69)</td></tr>
<tr><td rowspan="2">23. Styblo(35) (1967)<br><br>Czechoslovakia</td><td rowspan="2">• 15 years and older<br>• Resident of study district</td><td rowspan="2">General Community Survey</td><td rowspan="2">1961 - 1965</td><td rowspan="2">CXR with two blinded, independent reviewers</td><td rowspan="2">Microscopy of sputum and culture of sputum/laryngeal swabs on 1-3 occasions in all with symptoms or abnormal CXR and in a random selection not meeting these criteria</td><td rowspan="2">Clinical History</td><td rowspan="2">• Reviewed monthly if hospitalised<br>• If ambulatory, CXR annually ± bacteriology if symptomatic or CXR changes</td><td>cxr.neg | micro.neg | sympt.neg<br><br>to<br><br>cxr.unk | micro.pos | sympt.pos</td><td>• 17/73 000 (1-12)<br>• 33/73 000 (1-24)<br>• 49/73 000 (1-36)<br>• 66/73 000 (1-48)</td></tr>
<tr><td>cxr.neg | micro.neg | sympt.neg<br><br>to</td><td>• 102/73000 (1-12)</td></tr>
</table>

| | | | | | | | | | |
|---|---|---|---|---|---|---|---|---|---|
| | | | | | | | | cxr.unk \| micro.pos \| sympt.unk | • 123/73000 (1-24)<br>• 168/73000 (1-36)<br>• 175/73000 (1-48) |
| **24. Tuberculosis Society of Scotland(36, 37)** (1958, 1963)<br><br>SCOTLAND | • Age: >15 years<br>• European<br>• No bacilli on sputum smear or culture<br>• Abnormal CXR but of "doubtful activity" with no cavitation<br>• No known active extrapulmonary disease<br>• Not pregnant/ recently post-partum<br>• Not diabetic | Control/ Placebo arm | 1954 - 1959 | CXR | Sputum, gastric lavage or laryngeal swab for microscopy and/or culture | Authors noted that "chest radiograph was the only important sign of disease" | • 3-monthly reviews, including CXR, weight checks and ESR testing<br>• Follow up sputum testing was done in all those able to produce a sputum sample; laryngeal swabs or gastric lavage was only done if deterioration suspected | cxr.pos \| micro.neg \| sympt.neg<br><br>to<br><br>cxr.pos \| micro.pos \| sympt.unk | 9/95 (24) |

* Start and end points are labeled with three characteristics or states including a Chest Xray status (i.e. cxr.neg or cxr.pos or cxr.unk), a microbiological status (i.e. micro.neg, micro.pos, micro.unk, or micro.mixed), and a symptom status (i.e. sympt.neg, sympt.pos, sympt.unk or sympt.mixed). In these states, "neg" denotes negative, "pos" denotes positive, "unk" denotes unknown and "mixed" denotes a mixed group.
† The National Tuberculosis Association (NTA) developed criteria for assigning a clinical status to individuals with tuberculosis(38). These criteria and nomenclature changed over time but generally included groups such as "apparently cured", "arrested", "apparently arrested", "quiescent", and "active". The definition of these groups included radiographic, microbiological and clinical criteria over defined periods of time.
‡ This group were known to have been microbiologically positive on a prior occasion, then documented arrested disease, followed by a relapse

ATT=Antituberculosis Therapy; IUAT=International Union Against Tuberculosis; NTA=National Tuberculosis Association; UK=United Kingdom; USA=United States of America

### Supplementary Table 4: Descriptors on imaging assessments

| **Author** (Publication Year)<br><br>Country | Imaging Assessment | Descriptor/quoted text from study |
|---|---|---|
| **1. Alling** (1955)<br><br>USA | Inactive | "The clinical status of each person at the time of diagnosis was assessed on the basis of roentgenographic and bacteriologic data collected during the first six months following diagnosis…according to the Diagnostic Standards (1940 edition) published by the National Tuberculosis Association (NTA)."<br>"Arrested: For six months or more prior to the anniversary, the patient's sputum has been free of acid-fast bacilli and his serial chest roentgenograms have been compatible with stable disease. When more than a year has elapsed between clinic examinations and the chest roentgenograms show a stable lesion, the patient is continued in the arrested category throughout the interim." |
| **2. Anastasatu** (1985)<br><br>Romania | Inactive | "A group with an X-ray lesion extension smaller than 10cm$^2$ and with no epidemiological risk [and]… no "radiological activity" |
| **3. Aneja** (1979)<br><br>India | Active | "They had an abnormal shadow on a chest photofluorogram, read as pulmonary tuberculosis (TBP) by the Medical Officer"<br>"suspected to have active tuberculosis on X -ray evidence on single 70mm photofluorogram" |
| **4. Beeuwkes**<br>(1942)<br><br>USA | Mixed | "Latent (asymptomatic) lesions of first infection or childhood type were designated as calcified nodules, and latent infiltrations, childhood; latent lesions of reinfection or adult type, as latent apical tuberculosis."<br>"Latent (asymptomatic) infiltrations of either the childhood or adult type could be active (progressive or retrogressive), but they were classified as asymptomatic until the patient had symptoms, or physical signs of disease were demonstrated. At that time the diagnosis was changed to manifest (clinical) disease." |
| | Active | Manifest (clinical) tuberculosis, that which gives rise to physical signs or symptoms or both, was designated as childhood (first infection type) or adult (reinfection type) depending on the usual anatomical and clinical characteristics of the lesion |
| **5. Bobrowitz** (1947; 1949)<br><br>USA | Mixed | "Only patients with minimal tuberculosis according to the classification of the National Tuberculosis Association and with the characteristic roentgenological picture of pulmonary tuberculosis of the reinfection type were acceptable. These cases were further subdivided into four groups, according to the size of the infiltrations, In grouping the patients by type of lesion, we utilized the excellent definitions for the roentgenological character of the infiltration described by Reisner and Downes - Exudative, Productive and fibrotic, Exudative-productive, Fibro-calcific form" |
| 6. B**orgen** (1951;1952)<br><br>Norway | Active | "Cases submitted to control examination [were] grouped according to roentgenographic and clinical findings into 10 categories including:<br>Active non-destructive tuberculous pulmonary lesions - This group includes cases with pulmonary infiltrates which, according to the roentgenographic findings (soft, indistinct parenchymal lesions) and clinical findings (elevated temperature, increased sedimentation rate, loss of weight, etc.) were considered active lesions but where direct smears or culture failed to reveal tubercle bacilli in the sputum and after an observation period of 1.5-3.5 years no other etiology could reasonably be assumed." |
| | Inactive | "Inactive pulmonary lesions - This group includes cases with clear-cut, dense, streak-formed and, sometimes calcified lesions. In these cases the process showed no signs of activity during the observation period" |
| **7. Breu** (1954)<br><br>Germany | Mixed | "Bewusst beschränke ich mich bei der Aufzeigung der Entwicklungsreihen aus der geschlossenen in die bakteriologisch offene Tuberkulose, bzw. auf die angeführten röntgenologischen Erscheinungsformen (=Ia-b-Fälle der Fürsorgestatisik)…";"nur [...] als Ic/IIa angesprochenen Tuberkulosefälle [...], die sich später zu einer ansteckenden Lungentuberkulose entwickelten."<br>*Intentionally, in presenting the progression from closed to bacteriological open tuberculosis, meaning the roentgenological appearances (Ia-b-cases of the register); only as Ic/IIa classified cases that later progressed to a contagious tuberculosis. [According to old German tuberculosis classification, where Ia/b open, active on xray and contagious with or without bacteriology; Ic open, active but not contagious, with fuzzy appearance on xray; IIa closed, inactive with clear demarcation of lesion on xray]* |
| **8. Cowie** (1985) | Active | "New or enlarging apical lung lesions detected by routine 6-monthly chest radiography" |

| | | |
|---|---|---|
| South Africa | | |
| **9. Downes** (1938)<br><br>USA | Active<br>(Also micro pos) | Retrospectively captured imaging findings, based on clinical records |
| **10. Frimodt-Moller** (1965)<br><br>India | Active | Cases showing lung pathology on the survey films according to two independent readers were referred for bacteriological examination in the field with laryngeal swab cultures and sputum collection for microscopy and sputum culture as well as<br>for having another 70 mm X-ray film taken. The new film was compared with the former and cases thought to be of a non-tuberculous aetiology, or had only calcified lesions, were excluded. Other cases were ranked according to extent of the parenchymal lesion<br>The pretreatment films of the cases have for the purpose of this report been re-read and classified by the senior author with regard to extent of disease and cavitation according to the pattern of classification used by Fox and Sutherland (1956). The assessor did not know what treatment had been given or the fate of the individual cases studied. |
| **11. Hong Kong Chest Service** (1979(13), 1981(14), 1981(15), 1984(16))<br><br>HONG KONG | Active | “Patients admitted to the study…were diagnosed at the regular routine meetings of physicians of the Hong Kong Chest Service as having radiologically active pulmonary tuberculosis, not previously treated” |
| **12. IUAT Committee on Prophylaxis** (1982)<br><br>Europe | Inactive | “For the purpose of entry to the trial, fibrotic lesions were defined as well-delineated radiographic lesions of probable tuberculous origin, usually in the upper half of the lung, which had been stable during the year prior to entry” |
| **13. Lincoln** (1954)<br><br>USA | Inactive | “A review was made of all original diagnoses of minimal pulmonary tuberculosis made by the hospital staff during the years 1937 through 1947. In no case had a diagnosis of pulmonary tuberculosis been previously made”<br>“Two physicians, independently of one another and without knowledge of the subsequent clinical course, classified the diagnostic chest roentgenogram of each case according to the presence of pulmonary tuberculosis and the stage of disease. The classification of the stage of disease was based on the Diagnostic Standards (1940 edition), published by the National Tuberculosis Association (NTA). In the few cases in which there was disagreement, consultation was held to effect a mutually acceptable decision.”<br>“Arrested: For six months or more prior to the anniversary, the patient’s sputum has been free of acid-fast bacilli and his serial chest roentgenograms have been compatible with stable disease. When more than a year has elapsed between clinic examinations and the chest roentgenograms show a stable lesion, the patient is continued in the arrested category throughout the interim.” |
| **14. Manser** (1953)<br><br>Switzerland | Active<br>(Also micro pos) | Retrospectively captured imaging findings, based on clinical records |
| **15. Marshall** (1948)<br><br>UK | Active<br>(Also micro pos) | Study included patients with acute bilateral pulmonary tuberculosis of presumably recent origin, and patients who were unsuitable for collapse therapy. Old-standing disease and disease with thick-walled cavities excluded. Xrays were reviewed by a blinded radiologist. |
| **16. National Tuberculosis Institute** (1974; 1976; 1976; 1976; 1978; 1978; 1978; 1978; 1982)<br><br>India | Active | “Suggestive of tuberculous aetiology judged to be possibly, probably or definitely active tuberculous” |
| **17. Norregaard** (1990)<br><br>Denmark | Active | “The admission criteria were usually small X-ray shadows compatible with active pulmonary tuberculosis in smear negative patients. This evaluation was made at conference by the consultants…. All patients had infiltrates in the upper lobe of one or both lungs” |

| | | |
|---|---|---|
| **18. Okada** (2012)<br><br>Cambodia | Active | "A panel of experts composed of two respiratory physicians and/or radiologists made a decision on the final radiological findings. Based on the Japanese CXR classification for TB"<br>"A CXR suggestive of active TB was categorised as 'TB-suggestive CXR'" |
| **19. Orrego Puelma** (1945)<br><br>CHILE | Mixed | "Minimum or superficial lesions without apparent cavity, limited to a small area in one or both lungs. The total involvement, ignoring its distribution, should not exceed the equivalent in volume of the lung tissue which lies above the second chondrosternal junction and the spine of the fourth or body of the fifth thoracic vertebra on one side." "Furthermore, we studied only those cases which had two years' evolution, at least." |
| **20. Pamra** (1971)<br><br>India | Mixed | (a) No history of previous treatment for tuberculosis.<br>(b) No manifest tuberculous lesion in any organ other than the lungs.<br>(c) No evidence of diabetes or any other non-tuberculous disease.<br>(d) Apparent radiological stability of the lesions during the period of observation before inclusion in the study, this period being not less than 3 months or more than 3 years. |
| **21. Puffer** (1945)<br><br>USA | Mixed | "persons with lesions demonstrable by X-ray examination had no physical signs nor symptoms and gave no history of illness indicative of tuberculosis, they are classed as latent apical cases"<br>"The classification of lesions as minimal, moderately advanced and far advanced is in accordance with that recommended by the National Tuberculosis Association." |
| **21. Puffer** (1945)<br><br>USA | Inactive | "manifest cases were believed to be arrested at the time of first examination in this clinic."<br>"The classification of lesions as minimal, moderately advanced and far advanced is in accordance with that recommended by the National Tuberculosis Association." |
| **22. Sikand** (1959)<br><br>India | Inactive | Inactive Pulmonary Lesions<br>(i) Sputum negative by all methods or L. S. negative by culture;<br>(ii) Hardish shadows in the skiagrams;<br>(iii) No clinical or laboratory evidence suggestive of activity. |
| **22. Sikand** (1959)<br><br>India | Mixed | Lesions of Doubtful Activity<br>(i) Small soft infiltrations, stationary of regressive but not progressive;<br>(ii) Primary type of tuberculosis;<br>(iii) Known pulmonary cases under successful therapy. |
| **23. Styblo** (1967)<br><br>Czechoslovakia | N/A | Xray negative group |
| **24. Tuberculosis Society of Scotland** (1958, 1963)<br><br>SCOTLAND | Mixed | "Patients were accepted for the trial if the chest radiograph showed an abnormality considered to be tuberculous, this being the only important sign of disease"<br>"certain conditions also excluded acceptance- ... cavitation demonstrable on an ordinary posteroanterior radiograph"<br>"The clinicians were asked to classify their cases as 'acute' without evidence of fibrosis, or 'chronic', with fibrosis present." |

#### Supplementary Table 5: Descriptors of cohorts included in the Systematic Review

| | n (%) or median (IQR) |
|---|---|
| Cohort Types, n=34 | |
| • Clinical Cohort | 9 (24.5) |
| • Control arm of therapeutic intervention | 9 (24.5) |
| • General Community Survey | 7 (20.6) |
| • Household Contact Study | 4 (11.8) |
| • Occupational or Student Screening | 5 (14.7) |
| Study Location, n=34 | |
| • Africa | 1 (2.9) |
| • Asia | 11 (32.4) |
| • Europe | 10 (29.4) |
| • South America | 1 (2.9) |
| • North America | 11 (32.4) |
| Status at entry to cohorts, n=34 | |
| • Normal Chest X-ray/imaging | 4 (11.8) |
| • Abnormal Chest X-ray/imaging | 30 (88.2) |
| ○ Abnormal Chest X-ray/imaging, where the authors state that the extent of disease was active | 15 (44.1) |
| ○ Abnormal Chest X-ray/imaging, where the authors state that the extent of disease was inactive/fibrotic | 9 (26.5) |
| ○ Abnormal Chest X-ray/imaging, where the authors state that the extent of disease was "mixed" in this group or they did not describe/specify | 6 (17.6) |
| • Microbiological status† negative | 28 (82.4) |
| • Microbiological status† positive | 6 (17.6) |
| • Microbiological status† mixed or unknown | 0 (0.0) |
| • Symptomatic‡ | 7 (20.6) |
| • Asymptomatic‡ | 11 (32.4) |
| • Symptom status† mixed or unknown | 16 (47.1) |
| Demographics, n=34 | |
| • Sex distribution | |
| ○ Equal distribution of males and females | 5 (14.7) |
| ○ More females | 8 (23.5) |
| ○ More males | 11 (32.4) |
| ○ Unknown sex distribution | 10 (29.4) |
| • Age distribution | |
| ○ Minimum | 5 years |
| ○ Maximum | 83 years |
| ○ No details on age distribution specified | 10 (29.4) |
| Local Burden of TB Disease at time of cohort§, n=34 | |
| • Low | 0 (0.0) |
| • Mid | 2 (5.9) |
| • High | 9 (26.5) |
| • Not specified or unknown | 23 (67.6) |

* Studies were able to contribute ≥1 cohort
† To establish microbiological status, authors used a variety of methods which included sputum (spot or extended collection periods e.g. overnight), laryngeal swabs, and gastric aspirates
‡ Symptom status was as per author classification
§ As per World Health Organisation stratification

IQR = Interquartile Range; TB = Tuberculosis

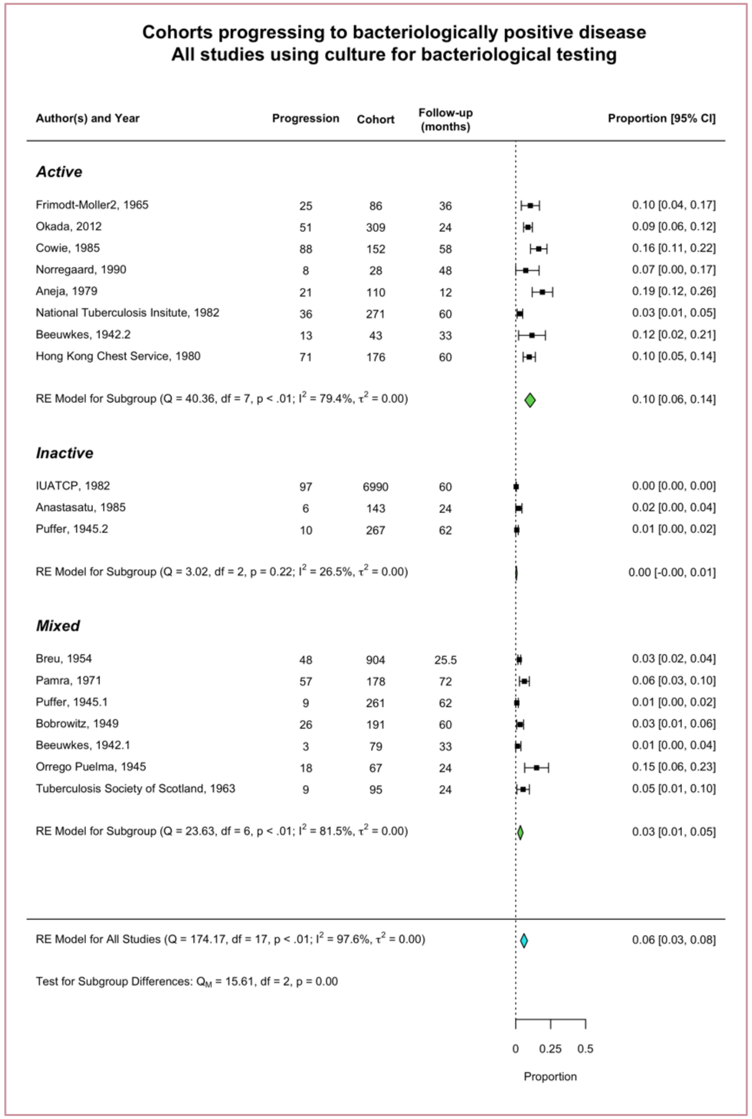

Supplementary Figure 1: Micro negative to positive, in those studies that used culture

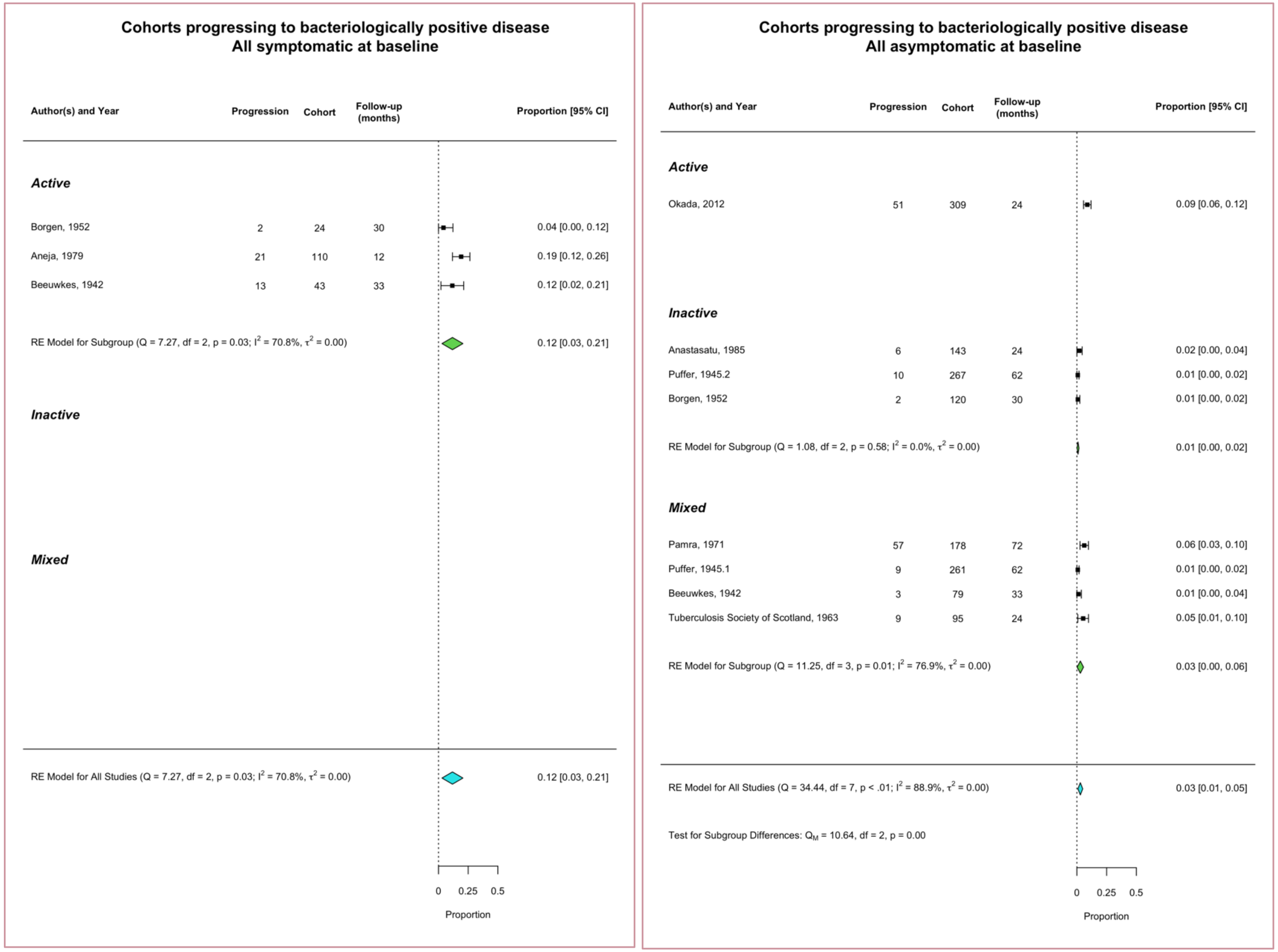

Supplementary Figure 2: Micro negative to positive, stratified by symptom and CXR status at baseline

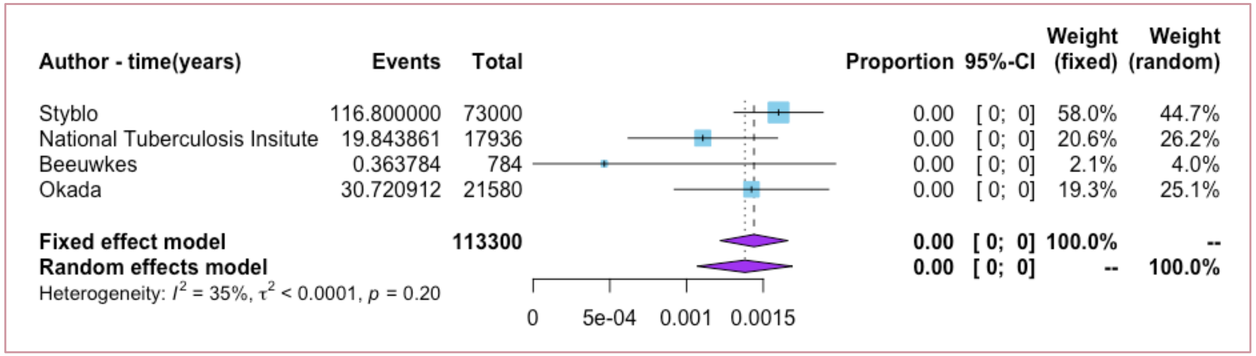

#### Supplementary Figure 3: CXR negative and Micro negative, to Micro positive

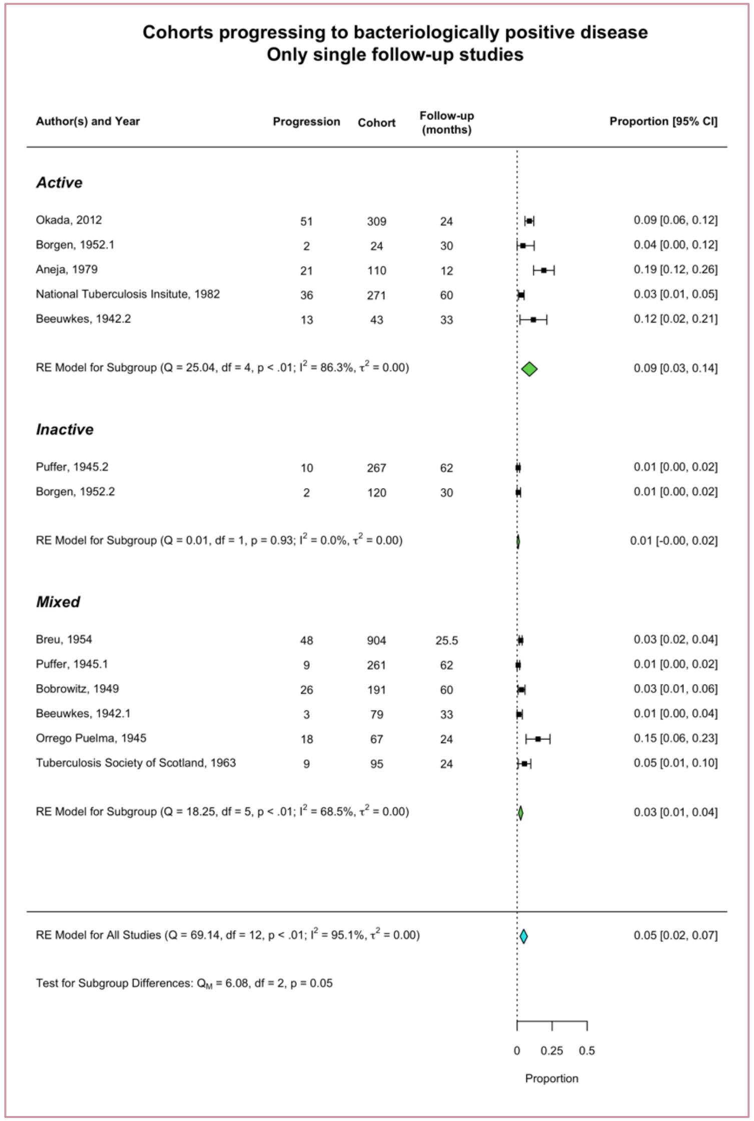

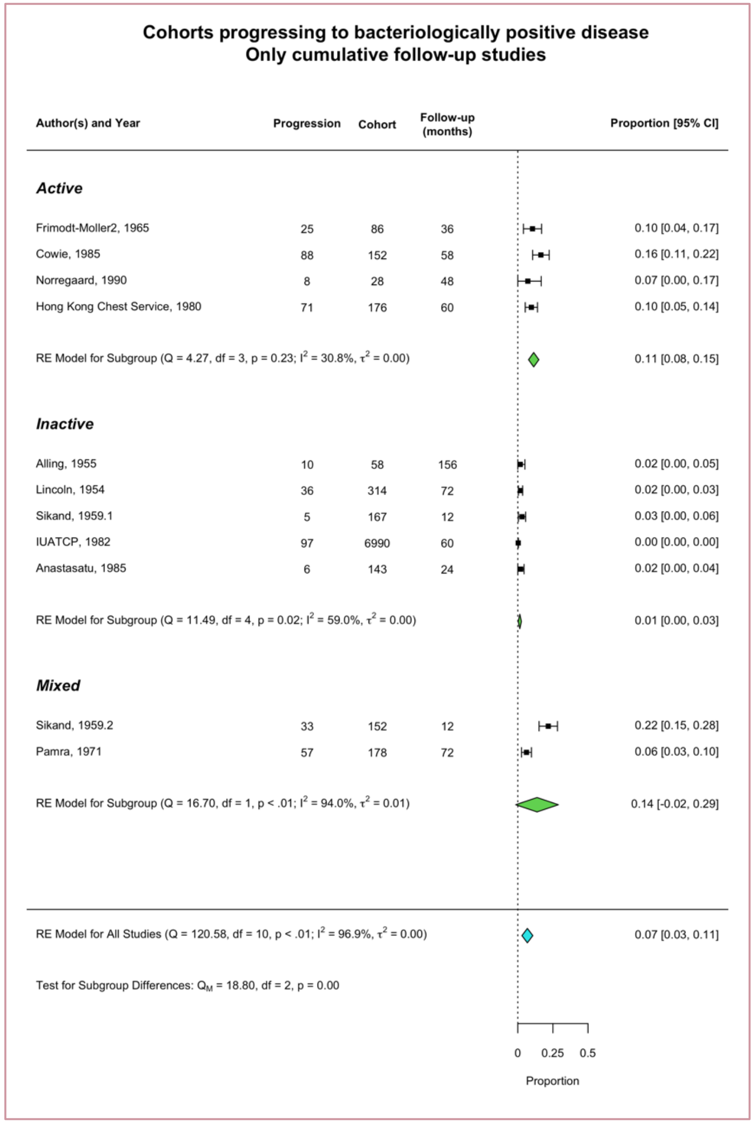

Supplementary Figure 4: Micro negative to positive, stratified by structure of follow up

#### Full Search Strategy: English

Filters to be applied within each:

- Languages: English, German & "Unknown language" (latter possible in Pubmed)
- Years: 1895 - 1960

a) Pubmed, Medline and Old Medline

| | Category | Search terms |
|---|---|---|
| **ENGLISH** | **Population:** | Pulmonary tuberculosis OR "Tuberculosis, Pulmonary"[Mesh] OR Incipient tuberculosis OR Phthisis OR Minimal tuberculosis OR Moderate tuberculosis OR Advanced tuberculosis |
| | **Intervention:** | Follow up studies OR Course OR Biological Evolution OR After-history OR Supervision OR "Early Diagnosis"[Mesh] OR "Epidemiological Study"[Mesh] OR Epidemiologic study OR Prognosis OR Prospective OR Longitudinal OR Roentgenographic Survey OR Radiography OR Observation OR Progression OR Progress OR Diagnosis |

((((((("tuberculosis, pulmonary"[MeSH Terms] OR ("tuberculosis"[All Fields] AND "pulmonary"[All Fields]) OR "pulmonary tuberculosis"[All Fields] OR ("pulmonary"[All Fields] AND "tuberculosis"[All Fields])) OR "Tuberculosis, Pulmonary"[Mesh]) OR (Incipient[All Fields] AND ("tuberculosis"[MeSH Terms] OR "tuberculosis"[All Fields]))) OR ("tuberculosis, pulmonary"[MeSH Terms] OR ("tuberculosis"[All Fields] AND "pulmonary"[All Fields]) OR "pulmonary tuberculosis"[All Fields] OR "phthisis"[All Fields])) OR (Minimal[All Fields] AND ("tuberculosis"[MeSH Terms] OR "tuberculosis"[All Fields]))) OR (Moderate[All Fields] AND ("tuberculosis"[MeSH Terms] OR "tuberculosis"[All Fields]))) OR (Advanced[All Fields] AND ("tuberculosis"[MeSH Terms] OR "tuberculosis"[All Fields]))) AND ((((((((((((((("follow-up studies"[MeSH Terms] OR ("follow-up"[All Fields] AND "studies"[All Fields]) OR "follow-up studies"[All Fields] OR ("follow"[All Fields] AND "up"[All Fields] AND "studies"[All Fields]) OR "follow up studies"[All Fields]) OR Course[All Fields]) OR ("biological evolution"[MeSH Terms] OR ("biological"[All Fields] AND "evolution"[All Fields]) OR "biological evolution"[All Fields])) OR After-history[All Fields]) OR ("organization and administration"[MeSH Terms] OR ("organization"[All Fields] AND "administration"[All Fields]) OR "organization and administration"[All Fields] OR "supervision"[All Fields])) OR "Early Diagnosis"[Mesh]) OR ("epidemiologic studies"[MeSH Terms] OR ("epidemiologic"[All Fields] AND "studies"[All Fields]) OR "epidemiologic studies"[All Fields] OR ("epidemiologic"[All Fields] AND "study"[All Fields]) OR "epidemiologic study"[All Fields])) OR ("prognosis"[MeSH Terms] OR "prognosis"[All Fields])) OR ("longitudinal studies"[MeSH Terms] OR ("longitudinal"[All Fields] AND "studies"[All Fields]) OR "longitudinal studies"[All Fields] OR "prospective"[All Fields])) OR Longitudinal[All Fields]) OR (Roentgenographic[All Fields] AND ("surveys and questionnaires"[MeSH Terms] OR ("surveys"[All Fields] AND "questionnaires"[All Fields]) OR "surveys and questionnaires"[All Fields] OR "survey"[All Fields]))) OR ("diagnostic imaging"[Subheading] OR ("diagnostic"[All Fields] AND "imaging"[All Fields]) OR "diagnostic imaging"[All Fields] OR "radiography"[All Fields] OR "radiography"[MeSH Terms])) OR ("observation"[MeSH Terms] OR "observation"[All Fields])) OR ("disease progression"[MeSH Terms] OR ("disease"[All Fields] AND "progression"[All Fields]) OR "disease progression"[All Fields] OR "progression"[All Fields])) OR Progress[All Fields]) OR ("diagnosis"[Subheading] OR "diagnosis"[All Fields] OR "diagnosis"[MeSH Terms])) AND (("1895/01/01"[PDAT] : "1960/12/31"[PDAT]) AND (Undetermined[lang] OR German[lang] OR English[lang]))

b) EMBASE and EMBASE CLASSIC

| | Category | Search terms |
|---|---|---|
| **ENGLISH** | **Population:** | “Pulmonary tuberculosis” OR “Incipient tuberculosis” OR Phthisis OR Minimal tuberculosis OR Moderate tuberculosis OR Advanced tuberculosis |
| | **Intervention:** | “Follow-up stud*” OR “Follow up stud*” OR Course OR “Biological Evolution” OR “After-history” OR Supervision OR Epidemiological Stud* OR Epidemiology OR Prognosis OR Prospective OR Longitudinal OR “Roentgenographic survey” OR Radiography OR Observation* OR Progress* OR Diagnosis |

c) Web of Science

| | Category | Search terms |
|---|---|---|
| **ENGLISH** | **Population:** | “Pulmonary tuberculosis” OR “Incipient tuberculosis” OR Phthisis OR Minimal tuberculosis OR Moderate tuberculosis OR Advanced tuberculosis |
| | **Intervention:** | “Follow up stud*” OR “Follow-up stud*” OR “Course” OR “Biological Evolution” OR “After-history” OR Supervision OR “Epidemiological Stud*” OR Epidemiology OR Prognosis OR Prospective OR Longitudinal OR “Roentgenographic Survey” OR Radiography OR Observation* OR Progress* OR Diagnosis |

#### Full Search Strategy: German

##### PUBMED

| | | |
|---|---|---|
| **GERMAN** | **Population:** | Pulmonale Tuberkulose OR Lungentuberkulose OR Lungen-Tuberkulose OR "Tuberkulose, pulmonal, pulmonale" [Mesh] OR anfangende Tuberkulose OR beginnende Tuberkulose OR Beginn der lungentuberkulose OR Fruehinfiltrat OR einsetzende Tuberkulose OR Schwindsucht<br><br>OR Phthisische Entiwicklung* OR Initialherd* OR Tuberculosis inappercepta OR Spitzenschwielen OR Spitzenherd* OR begrenzte* OR Tuberkulinreihenpruefungen OR Lungenschwindsucht OR Entstenhung OR Entwicklung* |
| | **Intervention:** | Längsschnittstudie OR Longitudinalstudie OR Verlaufsstudie OR Verlauf OR Entwicklung OR Verlauf OR Nachverfolgung OR Beobachtung OR Frühdiagnose OR Früherkennung OR "epidemiologische Studie" [Mesh] OR Prognose OR prospektiv OR vorausschauend OR longitudinal OR längsschnitt OR Röntgen OR röntgenographisch OR radiographisch OR radiologisch OR Beobachtung OR Betrachtung OR Folge OR Entwicklung OR Progression OR Fortschreitung OR Verlauf OR Fortschritt OR Diagnose |

##### EMBASE

| | | |
|---|---|---|
| **GERMAN** | **Population:** | "Pulmonale Tuberkulose" OR "Lungentuberkulose" OR "Lungen-Tuberkulose" OR "generalisierte Tuberkulose" OR "disseminierte Tuberkulose" OR "miliare Tuberkulose" OR "anfangende Tuberkulose" OR "beginnende Tuberkulose" OR "einsetzende Tuberkulose" OR Tuberk* OR Schwindsucht |
| | **Intervention:** | "Längsschnittstudie" OR "Längsschnitt-studie" OR "Longitudinalstudie" OR "Longitudinal-studie" OR "Verlaufsstudie" OR "Verlaufs-studie" OR Verlauf OR "Entwicklung" OR "Verlauf." OR "Nachverfolgung" OR Beobachtung OR epidemiologische Studie OR Epidemiologie OR Prognose OR Prospektiv OR Vorausschauend OR longitudinal OR längsschnitt OR "Röntgen" OR "röntgenographische Studie" OR "röntgenographische Untersuchung" OR "radiologische Studie" OR "radiologische Untersuchung" OR Radiologie OR Röntgen OR Beobachtung* OR Fortschritt* OR Diagnose |

##### Web of Science

| | | |
|---|---|---|
| **GERMAN** | **Population:** | "Pulmonale Tuberkulose" OR "Lungentuberkulose" OR "Lungen-Tuberkulose" OR "generalisierte Tuberkulose" OR "disseminierte Tuberkulose" OR "miliare Tuberkulose" OR "anfangende Tuberkulose" OR "beginnende Tuberkulose" OR "einsetzende Tuberkulose" OR Tuberk* OR Schwindsucht |
| | **Intervention:** | "Längsschnittstudie" OR "Längsschnitt- studie" OR "Longitudinalstudie" OR "Longitudinal-studie" OR "Verlaufsstudie" OR "Verlaufs-studie" OR "Verlauf" OR "Entwicklung" OR "Nachverfolgung" OR Beobachtung OR "epidemiologische Studie" OR Epidemiologie OR Prognose OR Prospektiv OR Vorausschauend OR longitudinal OR längsschnitt OR verlauf OR "Röntgen" OR "röntgenographische Studie" OR "röntgenographische Untersuchung" OR "radiologische Studie" OR "radiologische Untersuchung" OR Radiologie OR Röntgen OR Beobachtung* OR Fortschritt* OR Diagnose |

#### PRISMA Checklist(39)

| Section/topic | # | Checklist item | Reported on page # |
|---|---|---|---|
| **TITLE** | | | |
| Title | 1 | Identify the report as a systematic review, meta-analysis, or both. | 1 |
| **ABSTRACT** | | | |
| Structured summary | 2 | Provide a structured summary including, as applicable: background; objectives; data sources; study eligibility criteria, participants, and interventions; study appraisal and synthesis methods; results; limitations; conclusions and implications of key findings; systematic review registration number. | 3 |
| **INTRODUCTION** | | | |
| Rationale | 3 | Describe the rationale for the review in the context of what is already known. | 5-6 |
| Objectives | 4 | Provide an explicit statement of questions being addressed with reference to participants, interventions, comparisons, outcomes, and study design (PICOS). | 6,9 |
| **METHODS** | | | |
| Protocol and registration | 5 | Indicate if a review protocol exists, if and where it can be accessed (e.g., Web address), and, if available, provide registration information including registration number. | 9 |
| Eligibility criteria | 6 | Specify study characteristics (e.g., PICOS, length of follow-up) and report characteristics (e.g., years considered, language, publication status) used as criteria for eligibility, giving rationale. | 9 |
| Information sources | 7 | Describe all information sources (e.g., databases with dates of coverage, contact with study authors to identify additional studies) in the search and date last searched. | 9 |
| Search | 8 | Present full electronic search strategy for at least one database, including any limits used, such that it could be repeated. | 9 & Supplementary |
| Study selection | 9 | State the process for selecting studies (i.e., screening, eligibility, included in systematic review, and, if applicable, included in the meta-analysis). | 9-10 |
| Data collection process | 10 | Describe method of data extraction from reports (e.g., piloted forms, independently, in duplicate) and any processes for obtaining and confirming data from investigators. | 10-11 |

| | | | |
|---|---|---|---|
| Data items | 11 | List and define all variables for which data were sought (e.g., PICOS, funding sources) and any assumptions and simplifications made. | 11 |
| Risk of bias in individual studies | 12 | Describe methods used for assessing risk of bias of individual studies (including specification of whether this was done at the study or outcome level), and how this information is to be used in any data synthesis. | 10 |
| Summary measures | 13 | State the principal summary measures (e.g., risk ratio, difference in means). | 10-11 |
| Synthesis of results | 14 | Describe the methods of handling data and combining results of studies, if done, including measures of consistency (e.g., $I^2$) for each meta-analysis. | 12 |

| Section/topic | # | Checklist item | Reported on page # |
|---|---|---|---|
| Risk of bias across studies | 15 | Specify any assessment of risk of bias that may affect the cumulative evidence (e.g., publication bias, selective reporting within studies). | 27 |
| Additional analyses | 16 | Describe methods of additional analyses (e.g., sensitivity or subgroup analyses, meta-regression), if done, indicating which were pre-specified. | 11 |
| **RESULTS** | | | |
| Study selection | 17 | Give numbers of studies screened, assessed for eligibility, and included in the review, with reasons for exclusions at each stage, ideally with a flow diagram. | 13 (figure 2) |
| Study characteristics | 18 | For each study, present characteristics for which data were extracted (e.g., study size, PICOS, follow-up period) and provide the citations. | 16-18 & Supplementary |
| Risk of bias within studies | 19 | Present data on risk of bias of each study and, if available, any outcome level assessment (see item 12). | 14 |
| Results of individual studies | 20 | For all outcomes considered (benefits or harms), present, for each study: (a) simple summary data for each intervention group (b) effect estimates and confidence intervals, ideally with a forest plot. | Figure 3,4 & Supplementary |
| Synthesis of results | 21 | Present results of each meta-analysis done, including confidence intervals and measures of consistency. | Figure 3,4 & Supplementary |
| Risk of bias across studies | 22 | Present results of any assessment of risk of bias across studies (see Item 15). | N/A |
| Additional analysis | 23 | Give results of additional analyses, if done (e.g., sensitivity or subgroup analyses, meta-regression [see Item 16]). | Supplementary figures |
| **DISCUSSION** | | | |
| Summary of evidence | 24 | Summarize the main findings including the strength of evidence for each main outcome; consider their relevance to key groups (e.g., healthcare providers, users, and policy makers). | 25 |

| | | | |
|---|---|---|---|
| Limitations | 25 | Discuss limitations at study and outcome level (e.g., risk of bias), and at review-level (e.g., incomplete retrieval of identified research, reporting bias). | 26 |
| Conclusions | 26 | Provide a general interpretation of the results in the context of other evidence, and implications for future research. | 25-27 |
| **FUNDING** | | | |
| Funding | 27 | Describe sources of funding for the systematic review and other support (e.g., supply of data); role of funders for the systematic review. | 12 |

15. Hong Kong Chest Service. A controlled trial of 2-month, 3-month, and 12-month regimens of chemotherapy for sputum smear-negative pulmonary tuberculosis: the results at 30 months. 1981;

16. Hong Kong Chest Service, Centre TR. A Controlled Trial of 2-Month, 3-Month, and 12-Month Regimens of Chemotherapy for Sputum-Smear-Negative Pulmonary Tuberculosis: Results at 60 months. Am Rev Respir Dis. 1984;6.

30. Okada K, Onozaki I, Yamada N, Yoshiyama T, Miura T, Saint S, et al. Epidemiological impact of mass tuberculosis screening: a 2-year follow-up after a national prevalence survey. :7.
